# A Mitochondrial Supplement Improves Function and Mitochondrial Activity in Autism: A Double-Blind Place-bo-Controlled Cross-Over Trial

**DOI:** 10.1101/2025.01.13.25320480

**Authors:** Zoë Hill, Patrick J McCarty, Richard G. Boles, Richard E. Frye

## Abstract

Autism spectrum disorder (ASD) is associated with mitochondrial dysfunction but studies demonstrating the efficacy of treatments are scarce. We sought to determine whether a mitochondrial-targeted dietary supplement designed for children with ASD improved mitochondrial function and ASD symptomatology using a double-blind pla-cebo-controlled cross-over design. Sixteen children [mean age 9y 4m; 88% male] with non-syndromic ASD and mitochondrial enzyme abnormalities, as measured by MitoSwab (Religen, Plymouth Meeting, PA), received weight-adjusted SpectrumNeeds^®^ (NeuroNeeds, Old Lyme, CT) and QNeeds^®^ (NeuroNeeds) and placebos matched on taste, texture and appearance during two separate 12-week blocks. Which product received first was randomized. The treatment significantly normalized citrate synthase and complex IV activity as measured by the MitoSwab. Mitochondrial respiration of peripheral blood mononuclear cell respiration, as measured by the Seahorse XFe96 (Agilent, Santa Clara, CA) with the mitochondrial oxidative stress test, became more resilient to oxidative stress after the treatment, particularly in children with poor neurodevelopment. The mitochondrial supplement demonstrated significant improvement in standardized parent-rated scales in neurodevelopment, social withdrawal, and hyperactivity with large effect sizes (Cohen’s d’ = 0.77-1.25), while changes measured by the clinical and psychometric instruments were not significantly different. Adverse effects were minimal. This small study on children with ASD and mitochondrial abnormalities demonstrates that a simple, well-tolerated mitochondrial-targeted dietary supplement can improve mitochondrial physiology and ASD symptoms. Further larger controlled studies need to verify and extend these findings. These findings are significant as children with ASD have few other effective treatments.

## 1. Introduction

Autism spectrum disorder (ASD) is a behaviorally-defined disorder which a fects more than 2% of children in the United States with ASD affecting 4 times more males than females.^1^ Research has uncovered associated physiological abnormalities associated with ASD^2^ but high-quality clinical trials investigating biologically-targeted treat ents remain limited.^3^ Thus, the investigation of treatments which target underlying pathophysiological abnormalities and core and associated symptoms of ASD is urgently needed as there are few treatment options for children with ASD.^4^

Between 5%-80% of children with ASD demonstrate biomarkers of mitochondrial dysfunction while 5% obtain a diagnosis of mitochondrial disease.^5,6^ In comparison, mitochondrial disease affects less than 0.1% of the general population.^5,6^ Genetic defects involved in mitochondrial metabolism are rarely identified in children with ASD and co-morbid mitochondrial disease or mitochondrial dysfunction, and many reports describe moderate, rather than severe, deficiencies in electron transport chain (ETC) activity in those with ASD and mitochondrial disease.^5,6^ Perhaps more striking is that ETC activity in muscle, skin, buccal epithelium, fibroblasts and brain is significantly increased, rather than decreased, in some individuals with ASD.^5,6^ These reports suggest that abnormal mitochondrial function need not to be associated with genetic defects or depression of ETC function in individuals with ASD.

In our previous studies using the Seahorse 96eXF (Agilent, Santa Clara, CA) high-throughput respirometer our group has identified a subgroup of individuals with ASD with elevated mitochondrial respiration when analyzing both lymphoblastoid cell lines (LCLs) and peripheral blood mononuclear cells (PBMCs).^5^ These elevated respiratory rates were associated with a sharper decrease in ATP-associated respiration parameters (e.g. reserve capacity) as physiological stress was increased suggesting that the mitochondria were less resilient to physiological stress. To simulate physiological stress *in vitro* we developed the mitochondrial oxidative stress test (MOST) which systematically increases physiological redox stress using 2,3-dimethoxy-1,4-napthoquinone (DMNQ).^7,8^ The MOST assay can measure both overall mitochondrial respiration and resilience of the mitochondria to physiological stress.

Mitochondrial-targeted nutritional supplements have been studied in the general ASD population who do not specifically have mitochondrial dysfunction. Controlled studies have examined carnitine only.^9-12^ Open label and observational studies have examined carnitine,^13^ NAD and ribose,^14^ ubiquinol,^15^ ubiquinone^16^ and thiamine tetrahydrofurfuryl disulfide.^17^ Only one open-label trial treated 11 children with ASD and mitochondrial dysfunction with carnitine, coenzyme Q10 and alpha-lipoic acid.^18^ Three months of treatment improved mitochondrial function and behavior including the social withdrawal and inappropriate speech subscales from the Aberrant Behavior Checklist (ABC). The ABC and mitochondrial function worsened three months after withdrawal of the treatment. No blinded controlled trials have examined a mitochondrial targeted supplement on individuals with ASD who specifically have mitochondrial dysfunction.

Thus, we aim to determine whether a mitochondrial-targeted dietary supplement affects mitochondrial activity and ASD symptoms. Previous treatment studies were either open-label or did not measure mitochondrial activity. The primary aim of the study was to determine if a mitochondrial targeted treatment can normalize ETC activity as measured by the MitoSwab (Plymouth Meeting, PA) and increase mitochondrial resilience as measured by PBMC respiration. The secondary aim was to determine whether the treatment was associated with improvement in core ASD symptoms, behaviors, adaptive function and caregiver stress.

## 2. Methods

The study was approved by the Institutional Review Board at Phoenix Children’s Hospital (PCH; Phoenix, AZ) as #19-327 and WCG (Princeton, NJ) as #20224394, registered as NCT03835117 and conducted under Food and Drug Administration Center for Drug Evaluation and Research Division of Psychiatry IND 142751. Parents of participants provided written informed consent. This report is structured using the CONsolidated Standards Of Reporting Trials (CONSORT) guidelines.^19^ The CONSORT checklist can be found in supplementary materials and the CONSORT flow diagram is presented in Figure 1. No major changes were made to the methods or outcomes after commencement of the trial.

**Figure 1.**
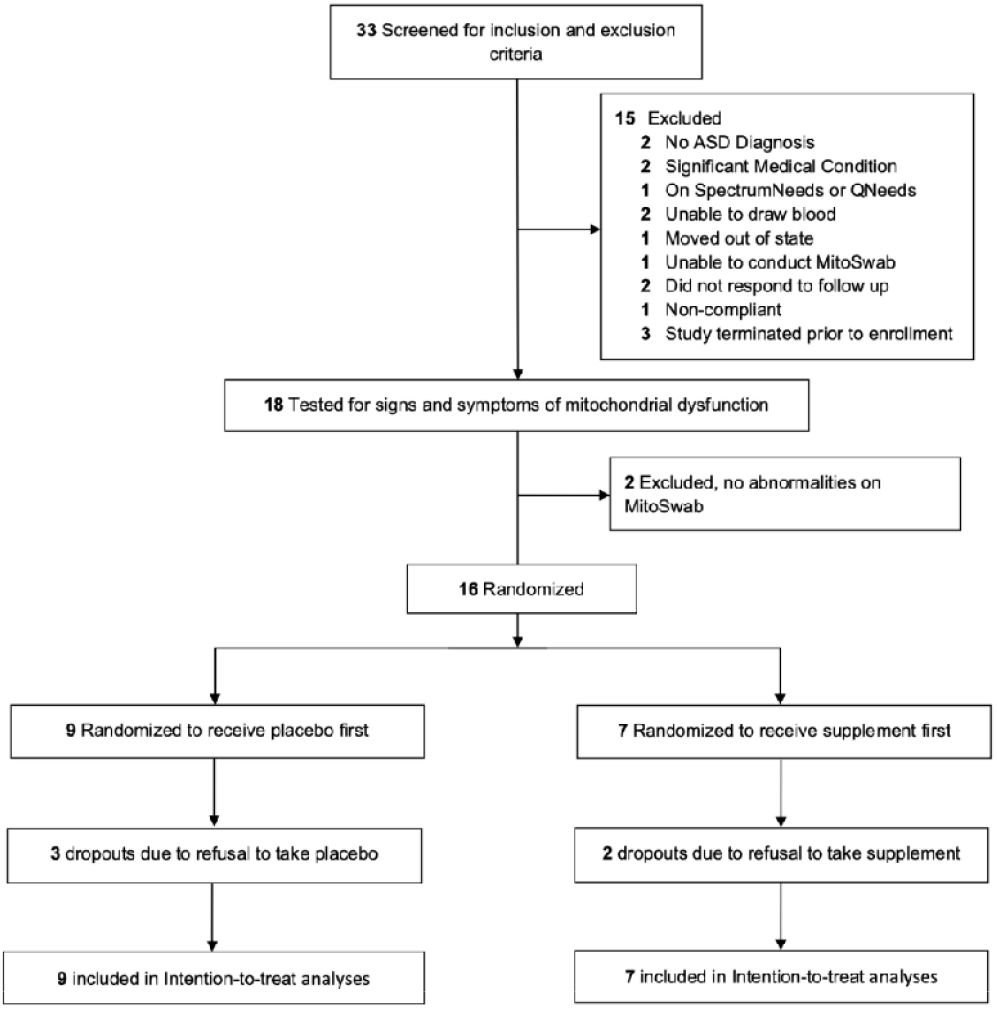
CONSORT Flow Diagram of Participants Through the Trial.

### 2.1. Study Design

This two-arm 24-week double-blind, randomized, placebo-controlled, cross-over study with a 1:1 allocation was performed at PCH (Phoenix, AZ) from February 01, 2022, to July 31, 2024.

### 2.2. Randomization

Randomization was performed using the National Cancer Institute Clinical Trial Randomization Tool with a maximally tolerated imbalance of 3. The research pharmacists had exclusive access to the randomization allocation.

### 2.3. Study Procedure

After initial assessments, participants were randomized to active or placebo treatment for 12 weeks. Assessments were repeated at the end of week 12 and then the participants received the opposite treatment that they received in the first 12 weeks. Assessments were again repeated after 12 weeks of treatment. Potential adverse effects (AEs), compliance and concomitant treatments were monitored at every visit and every 1 to 4 weeks. Families were compensated $50 for the screening, baseline, week 12 and week 24 visits.

### 2.4. Intervention

The active treatment was a weight-dosed combination of lemon flavored SpectrumNeeds^®^ and QNeeds^®^, both produced by NeuroNeeds (Old Lyme, CT; See Table 1), divided equally into two daily doses. SpectrumNeeds^®^ is a powder with 33 active ingredients that can be mixed into liquid or food. Thirty-one of the 33 supplements do not exceed the Tolerable Upper Intake Level (UL) set by the United States Institute of Medicine. The exceptions are:

**Table 1.** SpectrumNeeds^®^ and QNeeds^®^ Composition. According to the recommended daily serving size based on patient body weight, and each ingredient’s tolerable upper limit for children ages 4-8.

| Table 1. Spectrum Needs and QNeeds Composition. (ND = Not Defined) |  |  |  |  |  |  |
| --- | --- | --- | --- | --- | --- | --- |
| Daily Serving Size |  | 6.6g | 8.8g | 11g | 13.2g | Tolerable Upper Limit |
| Patient Body Weight | Units | 15kg-20kg | 21kg-40kg | 41kg-60kg | 61kg-100kg | Age 4-8 years |
| **SpectrumNeeds** |  |  |  |  |  |  |
| Vitamin C (ascorbic acid) | mg | 450 | 600 | 750 | 900 | ND |
| Vitamin D (cholecalciferol) | mcg | 11.25 | 15 | 18.75 | 22.5 | 75 |
| Vitamin E (d-α-tocopheryl acetate and mixed tocopherols) | mg | 37.5 | 50 | 62.5 | 75 | 300 |
| Thiamin (thiamin HCl) | mg | 18.75 | 25 | 31.25 | 37.5 | ND |
| Riboflavin | mg | 18.75 | 25 | 31.25 | 37.5 | ND |
| Niacinamide | mg | 18.75 | 25 | 31.25 | 37.5 | 15 |
| Vitamin B<sub>6</sub> (50% as pyridoxine HCl and 50% as pyridoxal-5-phosphate) | mg | 15 | 20 | 25 | 30 | 40 |
| (6S)-5-methyltetrahydrofolate | mcg | 1250 | 1667 | 2084 | 2500 | ND |
| Calcium folinate | mcg | 1250 | 1667 | 2084 | 2500 | 400 |
| Vitamin B<sub>12</sub> (as methylcobalamin) | mcg | 375 | 500 | 625 | 750 | ND |
| Biotin | mcg | 750 | 1000 | 1250 | 1500 | ND |
| Pantothenic acid (D-Ca pantothenate) | mg | 225 | 300 | 375 | 450 | ND |
| Calcium | mg | 45 | 60 | 75 | 90 | 2500 |
| Iodine (as K iodide) | mcg | 75 | 100 | 125 | 150 | 300 |
| Magnesium (Mg citrate and Mg malate) | mg | 150 | 200 | 250 | 300 | 110 |
| Zinc (Zn gluconate) | mg | 9 | 12 | 15 | 18 | 12 |
| Selenium (as selenomethionine) | mcg | 37.5 | 50 | 62.5 | 75 | 150 |
| Manganese (as Mn amino acid chelate) | mg | 0.75 | 1 | 1.25 | 1.5 | 3 |
| Chromium (Cr picolinate) | mcg | 37.5 | 50 | 62.5 | 75 | ND |
| Molybdenum (Na molybdate) | mcg | 75 | 100 | 125 | 150 | 600 |
| Potassium (K citrate) | mg | 37.5 | 50 | 62.5 | 75 |  |
| Creatine monohydrate | mg | 937.5 | 1250 | 1562.5 | 1875 |  |
| L-arginine | mg | 750 | 1000 | 1250 | 1500 |  |
| Carnitine blend (acetyl L-carnitine HCl and L-carnitine) | mg | 375 | 500 | 625 | 750 |  |
| Alpha ketoglutaric acid | mg | 225 | 300 | 375 | 450 |  |
| Ubiquinone | mg | 187.5 | 250 | 312.5 | 375 |  |
| Choline (choline bitartrate) | mg | 112.5 | 150 | 187.5 | 225 | 1000 |
| Inositol | mg | 75 | 100 | 125 | 150 |  |
| Alpha lipoic acid | mg | 75 | 100 | 125 | 150 |  |
| D-beta, D-gamma, D-delta tocopherols | mg | 3.3 | 4.4 | 5.5 | 6.6 | 300 |
| **QNeeds** |  |  |  |  |  |  |
| Co-enzyme Q10 (Ubiquinol) | mg | 200 | 300 | 400 | 400 |  |

1. Niacin is at about the UL. Niacinamide (also known as nicotinamide), the form of niacin in SpectrumNeeds^®^, does not cause flushing.
2. Folate exceeds the UL. However, the UL only applied to synthetic folate, also known as folic acid. There is no known UL for non-synthetic folate, which is the type of folate used in SpectrumNeeds^®^. Half of the total folate is microbial sourced Quatrefolic™ (6S)-5-Methyltetrahydrofolic acid Glucosamine salt. Half is calcium folinate, which is about one-half the dose recommended for pregnant women with a prior child born with a neural tube defect and one half to one-twenty-fifth the dosing given to children with ASD and suspected cerebral folate deficiency. These dosages are known to be generally well tolerated in ASD children.^20^

Ubiquinol (reduced form of coenzyme Q10, the principal electron carrier in the respiratory chain) is provided as a QNeeds^®^ gel capsule. SpectrumNeeds^®^ could be mixed in any liquid, and the QNeeds^®^ gel capsule could be opened and the liquid squeezed out into any liquid or food.

Certificate of analysis was provided by an independent analytical service (Eurofins USA, Petaluma CA) with a verification of a potency of at least 99% for every active ingredient, and undetectable toxic or biological (bacteria, yeast, or mold) contamination.

Placebos had the same look, feel and taste as the active treatments. SpectrumNeeds^®^ is a silica powder flavored by monk fruit extract, so the inert silica powder with the same favoring was used in the control. Three lemon-flavored placebos were produced for taste testing. Ten staff members, some familiar with SpectrumNeeds^®^, were given each placebo paired with the supplement to taste head-to-head. The placebo, which was most indistinguishable, as represented by a 50% choice of the placebo or supplement, was used for the trial. This was also performed by looking at the powder color. In QNeeds^®^ the ubiquinol is carried by purified water and glycerin colored by caramel liquid, so for the placebo of the same carrier and color was used in the gel capsule without ubiquinol.

Parents were asked about missed doses and returned containers were examined for adherence, which was calculated by the research pharmacy.

### 2.5. Inclusion and Exclusion Criteria

Participants were recruited from the senior author’s ASD clinic at PCH. Inclusion criteria included: (i) a diagnosis of ASD, (ii) age 2 years 6 months to 17 years 3 months years of age; (iii) weight 15 kg to 100 kg; (iv) Childhood Autism Rating Scale, 2^nd^ Edition (CARS-2) score meeting criteria for ASD; (v) general cognitive ability (GCA) ≥ 30 on the Differential Abilities Scale (DAS); (vi) unchanged complementary, traditional, behavioral and education therapy 8 weeks prior to enrollment; (vii) intention to maintain ongoing therapies constant throughout the trial; and (ix) abnormal mitochondrial activity as assessed by the buccal swab enzymology testing.

Exclusion criteria included: (i) serious behavioral problems (tantrums, aggression, self-injury) for which another treatment is warranted; (ii) significant medical condition that would be incompatible with the treatment; (iii) taking anticonvulsant medication for seizures or active epilepsy; (iv) diagnosis of Mitochondrial Disease; (v) current use of SpectrumNeeds^®^ or QNeeds^®^ or an equivalent mitochondrial supplement. Participants who were on standard multivitamins were allowed to participate in the study since the vitamin doses in such multivitamins are far below that of SpectrumNeeds. Those that were on high dose (defined as twice or more of the RDA) of three or more of the components of SpectrumNeeds or QNeeds were excluded.

The ASD diagnosis was confirmed using the lifetime version of the Autism Diagnostic Interview-Revised by an independent research reliable rater.

### 2.6. Behavioral and Developmental Outcome Measures

#### 2.6.1. Clinician Rated Measures

##### Childhood Autism Rating Scale, 2^nd^ Edition (CARS)

The CARS can both diagnose ASD and assess ASD severity and is conducted by a certified evaluator who both interacts with the participant and parent. The reliability is 0.90.^21^ Both the Standard and the High-Functioning versions were used as applicable. Clinical Global Impression - Severity (CGI-S): This is a 7-item scale ranging from a score of 1 for “Normal” to 7 for “Extreme.”^22^

##### Clinical Global Impression for Improvement (CGI-I)

The CGI-I is a 7-point measure of overall symptomatic change compared to baseline.^22^ Scores range from 1 (“very much improved”) through 4 (“unchanged”) to 7 (“very much worse”).

##### Children’s Yale-Brown Obsessive-Compulsive Scales-ASD (CYBOCS-ASD)

The CYBOCS-ASD is a modified version of the CYBOCS developed for children with obsessive-compulsive disorder^23^ and has established reliability and validity^24^ and is sensitive to change.^25^

##### Ohio State University Clinical Impressions Scale (OACIS)

The OACIS is a clinical rating of severity and improvement, using the CGI paradigm, for 10 ASD-related symptoms, which has good inter-rater and cross-cultural reliability.^26^ The rating is derived from an interview with the parent and observations and interactions with the patient.

#### 2.6.2. Psychometrician Rated Scales

##### Differential Abilities Scale (DAS)

This scale assesses cognitive abilities and early learning and has been validated against other cognitive ability scales (Wechsler, Bayley, etc.). The CGA index provides a measure similar to an intellectual quotient.

#### 2.6.3. Parent Rated Scales

##### Vineland Adaptive Behavior Scale (VABS) III

The Vineland Adaptive Behavior Scale (VABS) III is a widely used standardized, well-validated assessment tool for children with developmental delays that measures functional abilities.^27^ It is a valid measure of social impairments in children with ASD.^28^ The VABS relies on an informant (caretaker) to complete.

##### Aberrant Behavior Checklist (ABC)

The ABC measure disruptive behavior and has convergent and divergent validity.^29^

##### Caregiver Strain Questionnaire (CGSQ)-Short Form

The CGSQ is a 7-item parent self-report questionnaire that measures the burdens associated with raising a child with special needs.

##### Parent Rated Autism Symptomatic Change Scale (PRASC)

The PRASC is a 12-item questionnaire that asks parent to rate changes in symptoms from “much better” to “much worse” on a 7-point Likert scale compared to before the intervention.^30^ The parents are asked to complete this scale every 4 weeks.

##### Parent-Rated Anxiety Scale for Youth with Autism Spectrum Disorder (PRAS-ASD)

The PRAS-ASD assesses anxiety in ASD with excellent test-retest reliability and validity.^31^

Sensory Profile 2 (SP2): The SP2 measures sensory processing difficulties in the areas of auditory, visual, touch, movement, body position and oral sensation. The SP2 has been found to correlate well with other common instruments and have good reliability.

### 2.7. Establishment and Maintenance of Assessment Fidelity

Research staff were trained and certified by a licensed psychologist prior to performing assessments. During the trial a psychologist supervised staff and provided feedback and retraining, if necessary.

### 2.8. Adverse Effect Monitoring

Common medical symptoms were monitored at every visit using the modified Dosage Record Treatment Emergent Symptom (MDOTES) scale, a 34-item form which specifically queries common symptoms of major body systems, activity, sleep, appetite, and general health. At baseline, the severity of any ongoing symptoms was obtained. New symptoms or pre-existing symptoms that significantly worsened were classified as AEs and rated as mild, moderate or severe, and are counted at the highest level of severity. AEs were considered related to the treatment if they started or significantly worsened following the start of the trial. If AEs were persistent or severe, the parents were offered the option of halving the dose or discontinuing the intervention. The dose could only be reduced once and was never increased if reduced.

Suicidality was assessed using the Columbia-Suicide Severity Rating Scale (C-SSRS) and the endorsement of self-injury items on the ABC.

### 2.9. Measures of Mitochondrial Enzymology

We measured the activity of three ETC complexes using a validated buccal swab procedure.^32^ Buccal cells were collected using four Catch-All Buccal Collection Swabs (Epicentre Biotechnologies, Madison, WI) by firmly pressing against the inner cheek while twirling for 30s. Swabs were clipped, placed in 1.5ml microcentrifuge tubes and transported to the laboratory on dry ice where they are then stored at -80°C.

Buccal extracts were prepared using an ice-cold buffered solution (Buffer A, ABCAM) containing protease inhibitor cocktail and membrane solubilizing non-ionic detergent and cleared of insoluble cellular material by high-speed centrifugation at 4°C. Duplicate aliquots of the protein extract were analyzed for protein concentration using the bicinchoninic acid method (Pierce Biotechnology, Rockford, IL).

Dipstick immunocapture assays measured ETC complex I (C1) activity using 50µg extracted protein.^32-34^ Signals were quantified using a Hamamatsu Immunochromato Reader (MS 1000 Dipstick Reader). Raw mABS (milliAbsorbance) results were corrected for protein concentration. ETC complex IV (C4) and citrate synthase (CS) activity were assessed using standard spectrophotometric procedures in 0.5ml reaction volume.

ETC complex II (C2) was measured using extract prepared from 1-2 buccal swabs in ice-cold 300ul STE buffer containing lauryl maltoside and a protease inhibitor cocktail and vortexed for 1min. After centrifugation of the extract at 15,000g for 8min, the supernatant was used. C2 activity was spectrophotometrically determined in succinate-driven reactions by measuring the ubiquinone-initiated oxidation of DCPIP as gauged by the decrease in absorbance at 600nm.^35^

Specific activities of respiratory complexes were initially expressed as nanomoles per mg protein per minute and normalized to standardized values (z-scores) using the control mean and standard deviation to better represent the deviation of mitochondrial function from normal.

### 2.10. Measures of Mitochondrial Respiration

#### Blood Collection and Processing

Participants underwent a morning fasting blood draw at screening, and at the 12-week and 24-week visits. Samples were centrifuged at 1500g for 10 mins at 4°C to separate plasma. Plasma was removed and replaced with room temperature wash buffer containing Ca^+2^/Mg^+2^-free PBS with 0.1% bovine serum albumin and 2mM EDTA. Diluted blood was then layered on top of Histopaque-1077 (Sigma Aldrich, St. Louis, MO, USA) and centrifuged at 400g for 30mins at room temperature. PBMCs were removed from the interface and were washed twice with wash buffer. The viable cells were counted using trypan blue exclusion and a Countess II Automated Cell Counter (Invitrogen, ThermoFisher Scientific, Waltham, MA). The isolation procedure duration was 90-120mins in order to perform the Seahorse XFe96 assay within the optimal timeframe.^36^

#### Mitochondrial Respiration Assay

PBMCs were placed in assay media (unbuffered RPMI without phenol red) supplemented with 1mM pyruvate, 2mM glutamate and 25mM glucose warmed to 37°C and pH adjusted to 7.4 prior to cell suspension. XFe96 plates (Agilent Technologies, Santa Clara, CA) were prepared by adding 25µL of 50µg/mL poly-D-lysine (EMD Millipore, Burlington, MA) for two hours, washing with 250µL sterile water and drying in a laminar flow hood overnight prior to seeding with 4 x 10^5^ viable PBMCs per well. After seeding, the plates were spun with slow acceleration (4 on a scale of 9) to a maximum of 100g for 2mins and allowed to stop without braking (Eppendorf Model 5810R Centrifuge, ThermoFisher Scientific). The plate orientation was reversed and spun again in the same fashion. Prior to the Seahorse assay, XFe96 wells were visualized using an inverted microscope to ensure PBMCs were evenly distributed in a single layer. At least four replicate wells were used to improve assay reliability. Runs with clear measurement probe failure, reagent injection failures, or non-physiology measurements (ATP-linked respiration (ALR) or proton leak respiration (PLR) < -1 pmol/min) were eliminated.

The Seahorse measures oxygen consumption rate (OCR) in real-time using a 96-well plate.^37,38^ We have linked variation in mitochondrial function in PBMCs to immune abnormalities^39^, prenatal environmental exposures^40,41^ and neurodevelopmental regression^7^ and have shown excellent intraclass correlation for our PMBCs assay.^42^ The Seahorse assay is a 4-step process which monitors OCR three times during four distinct periods of time in response to various reagents that activate or inhibit the ETC (See Supplementary Figure 1) from which several key parameters are derived.

#### Redox Challenge

PBMCs were exposed to multiple concentrations of DMNQ (Sigma-Aldrich, St. Louis, MO, USA) at 37 °C in a non-CO_2_ incubator for 1h prior to the Seahorse assay.^7^ DMNQ generates both superoxide and hydrogen peroxide similar to levels generated by nicotinamide adenine dinucleotide phosphate oxidase *in vivo*.^43^ DMNQ (5mg/mL) was diluted in DMEM XF assay media (Agilent Technologies) into a 10X stock and added to cells in an XF-PS plate.^8^ We have verified that DMNQ increases superoxide in PBMCs.^7^

### 2.11. Statistical Analysis

An intention-to-treat analysis^44^ is the gold standard for clinical trials due to its powerful non-biased analysis.^45^ The analysis was performed using PASW Statistics version 28.0.0.0 (IBM SPSS Statistics, Armonk, NY), and graphs were produced using Excel version 14.0 (Microsoft Corp, Redmond, WA). A two-tailed alpha of 5% was used as a cutoff for significance.

Multiple imputation (50) was conducted for missing data since data were lacking at random.^44,46^ Sensitivity analysis checked for systematic bias.^47^ Original and processed data were not significantly different (p > 0.05).

Mixed-effects regression models^48^ estimated the effect and effect size of the treatment. The models included the effect of time and a random intercept to account for individual subject mean and variation. The models tested the *a priori* hypothesis that the change in the outcome measure was different for the active treatment as compared with the placebo treatment by calculating the interaction between treatment and time using a F-test with a p<0.05. With the current N=16, for a medium/large effect size, f=0.25/0.40, with an alpha of 0.05, repeated measures F-test with two groups and two measurements, correlation among repeated measures of 0.8 and non-sphericity correction = 1.0, the power is 84%/99%.

For the Seahorse mitochondrial measurements, the effect of DMNQ was modeled as a polynomial. Additionally, analyses were conducted on the Seahorse data divided into two equal sized high functioning and low functioning subgroups as defined as above or below the mean VABS Adaptive Behavioral Composite (VABC) score.

A responder analysis was conducted using mixed-model logistic regression. Responders were defined as those that exceeded the minimal clinical important difference (MCID), if it was available. The MCID has been defined for ABC,^3^ VABS,^3^ CYBOCS-ASD^49^ and CARS.^50^ For CGI-I and OACIS improvement scales, a rating of ‘much better’ or ‘very much better’ is the standard for defining response. Age, baseline language and baseline overall development (as indexed by the VABC Standardized Score) were entered as potential covariates.

The frequency of AEs by severity was evaluated using a Fisher’s exact test.

## 3. Results

### 3.1. Participants

Thirty-three participants were screened with 17 failing screenings (Figure 1). Nine participants [Mean (SD) Age 9y 4m (5y 1m); 88% male] were randomized to receive the placebo first (group A) and seven were randomized to receive the treatment first (group B). None of the participants had been diagnosed with genetic disorders, half experienced a neurodevelopmental regression during development, and one had seizures (Supplementary Table 1). Compliance was excellent (>90%) although the placebo supplement powder had slightly non-significantly different lower compliance (80%). Most participants demonstrated a significant elevation in citrate synthase and a deficit in C1. Characteristics were not significantly different across groups.

### 3.2. Mitochondrial Enzymology

CS was elevated prior to treatment and then significantly decreased toward normal with the supplement treatment in most of the participants (Figure 2A) [F(1,24)=6.49, p<0.05].

**Figure 2.**
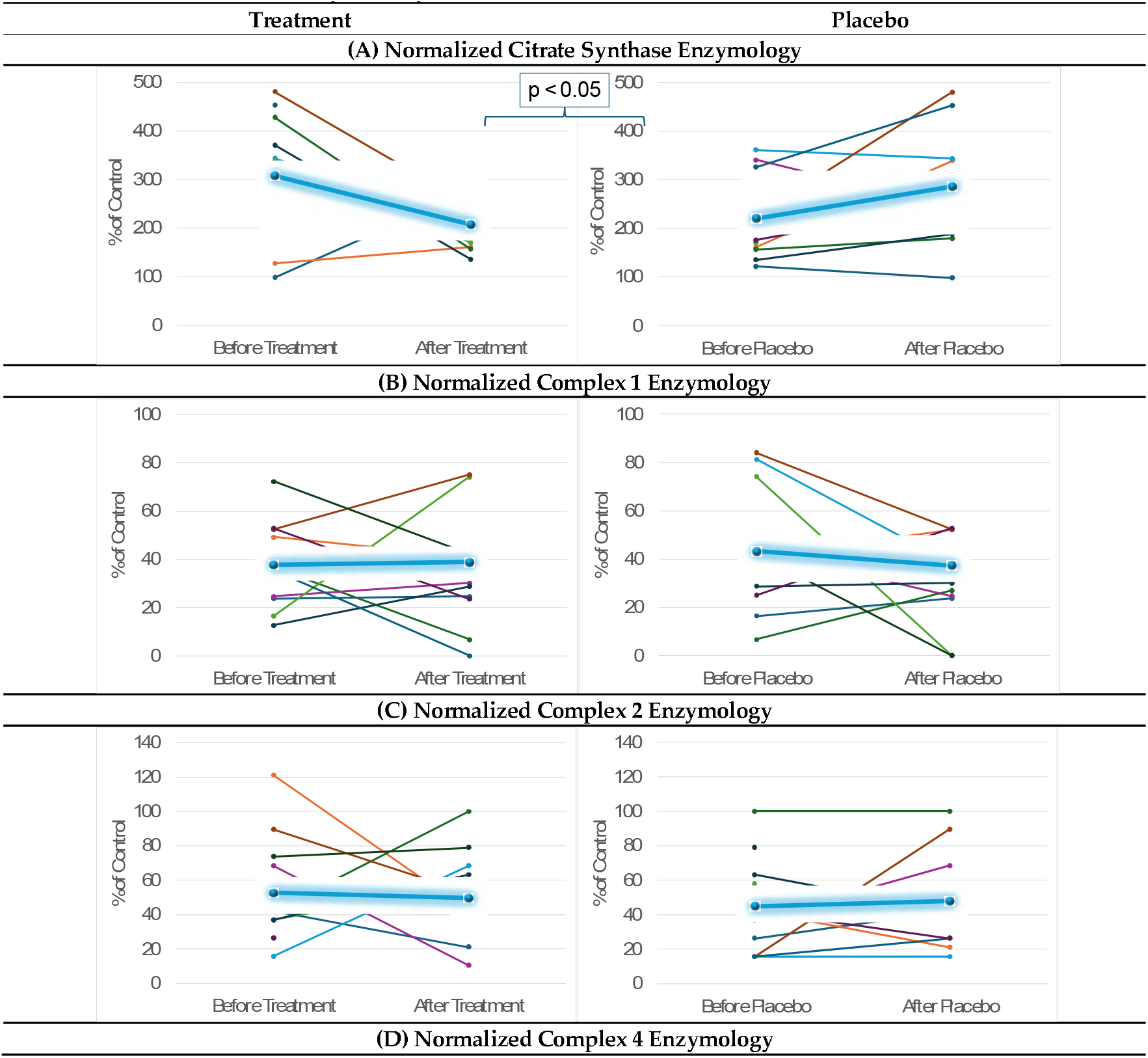
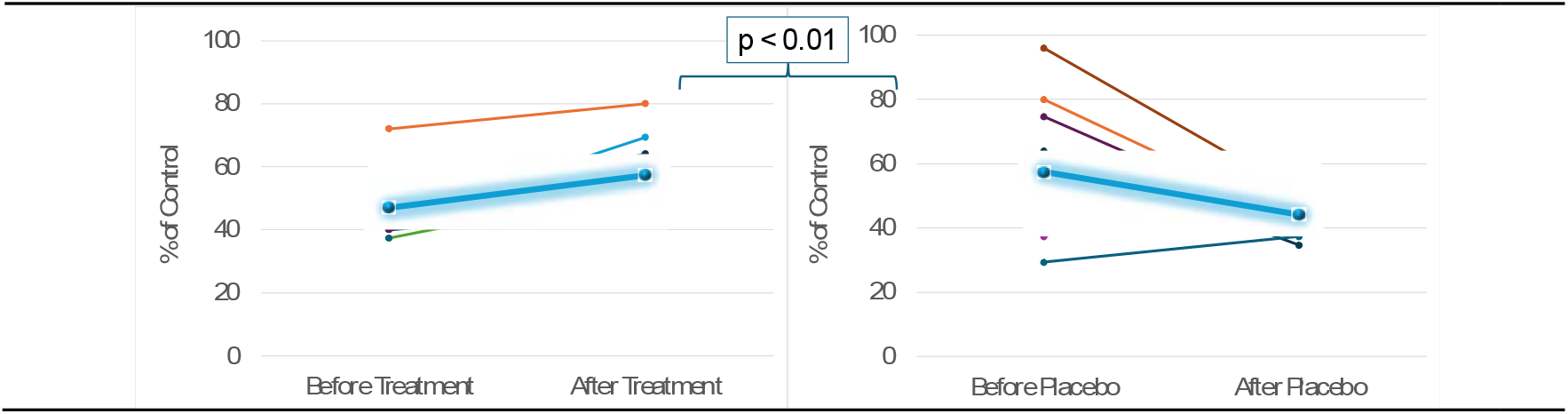
Mitochondrial Function. Function was measured over the course of 12 weeks before and after treatment and placebo. The highlighted blue lines represent the average effect for the group. Mixed-model regression was used to control for the repeated subjects level effects while determining whether the effect of treatment vs placebo affected the change over time.

Corrected (Figure 2B,C) Complex 1 and 2 activity was not significantly influenced by mitochondrial supplementation. Complex 4 activity corrected for CS [F(1,23.9)=8.28, p<0.01; See Figure 2D] significantly increased with mitochondrial supplementation as compared to placebo.

### 3.3. Mitochondrial Respirometry

The supplement increased the average ATP-linked respiration (ALR) [treatment x visit: F(1,1515)=3.99,p<0.05, Figure 3A] and prevented ALR from decreasing as physiological stress increased [treatment x visit x DMNQ: F(1,1506)=12.18, p<0.001, Figure 3A], demonstrating improved mitochondrial resilience when the supplement was taken. In comparison, ALR decreased with physiological stress with the placebo treatment.

**Figure 3.**
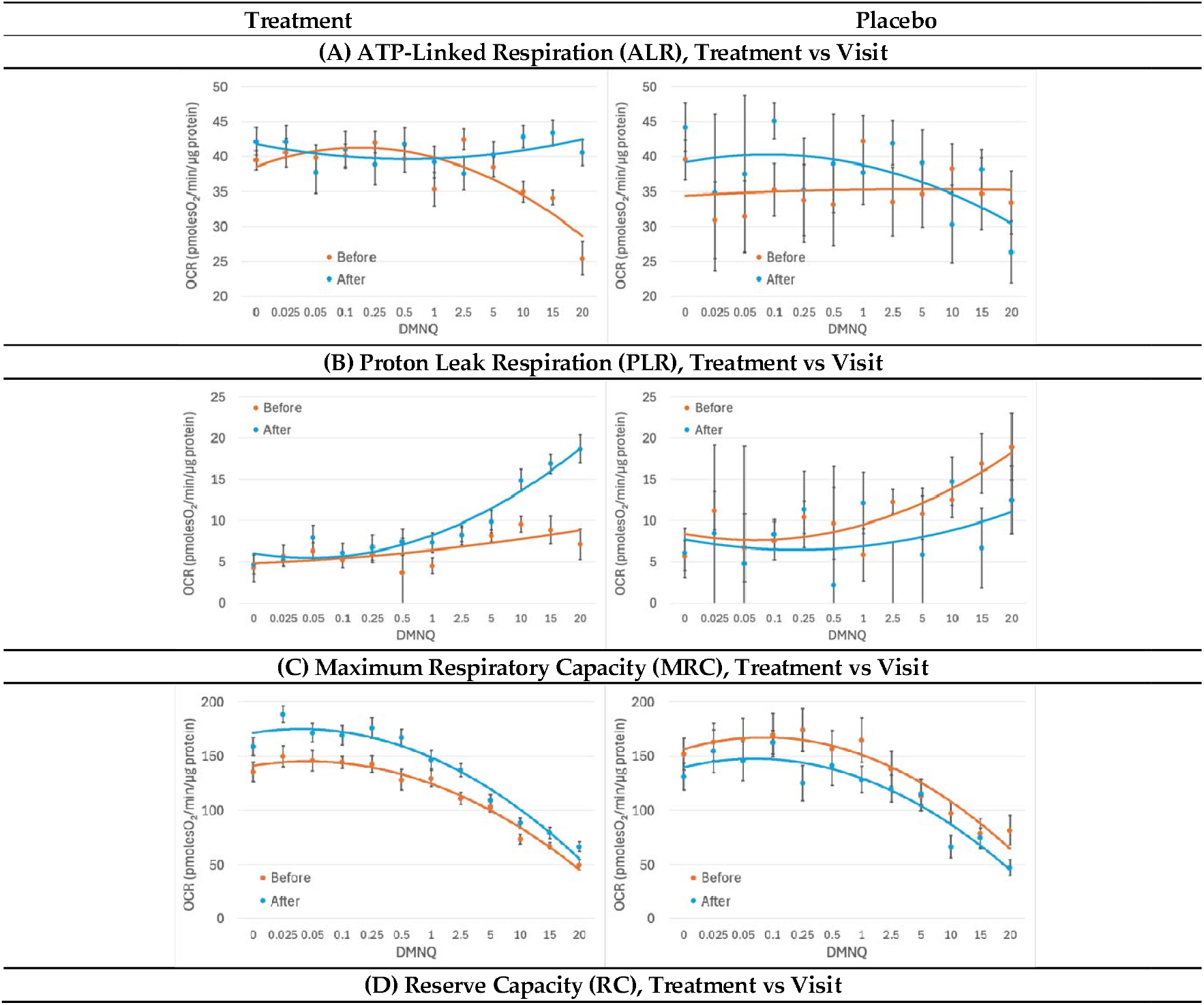
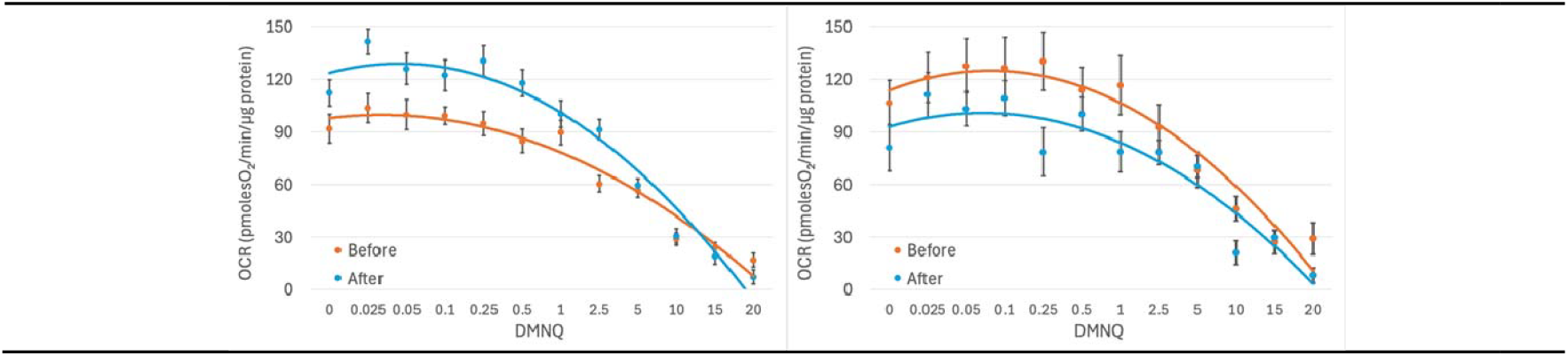
Mitochondria Effects. Effects were measured over the course of 12 weeks before and after treatment and placebo. Standard error bars are standard error of the mean. Mixed-model regression was used to control for the repeated subjects level effects while determining whether the effect of treatment vs placebo affected the change over time. Oxygen consumption rate (OCR); µM 2,3-dimethoxy-1,4-napthoquinone (DMNQ).

After treatment with the supplement, PLR increased much more as physiological stress was increased [treatment x visit x DMNQ: F(1,1514)=4.41,p<0.05, Figure 3B], suggesting a greater use of PLR to control physiological stress when the supplement was taken. In contrast the opposite effect occurred with the placebo.

Treatment with the supplement significantly increased the average maximum respiratory capacity (MRC) [treatment x visit: F(1,1508)=66.07,p<0.001, Figure 3C], demonstrating that the mitochondrial-targeted supplements improved overall mitochondrial capacity as compared to placebo. MRC did not statistically significantly change with the placebo treatment.

The mitochondrial supplement increased the average reserve capacity (RC) [treatment x visit: F(1,1511)=97.61,p<0.001] and change in RC with increasing physiological stress [treatment x visit x DMNQ: F(1,1492)=20.28,p<0.001, Figure 3D], increasing the average RC for low and moderate levels of physiological stress. RC did not statistically significantly change with the placebo treatment.

RC after the treatment dipped below the RC before treatment for high physiological stress levels. This suggests that the mitochondrial-targeted supplements improved mitochondrial resistance to mild and moderate physiological stress but lost this ability at high levels of physiological stress.

We investigated whether the effect of the supplement was dependent on the level of developmental impairment, as defined by the VABC. For all the respiratory parameters except PLR, the effect of the supplement was more pronounced in the low functioning group (Figure 4, graphs for the high-functioning subgroup can be found as Supplemental Figure 2).

**Figure 4.**
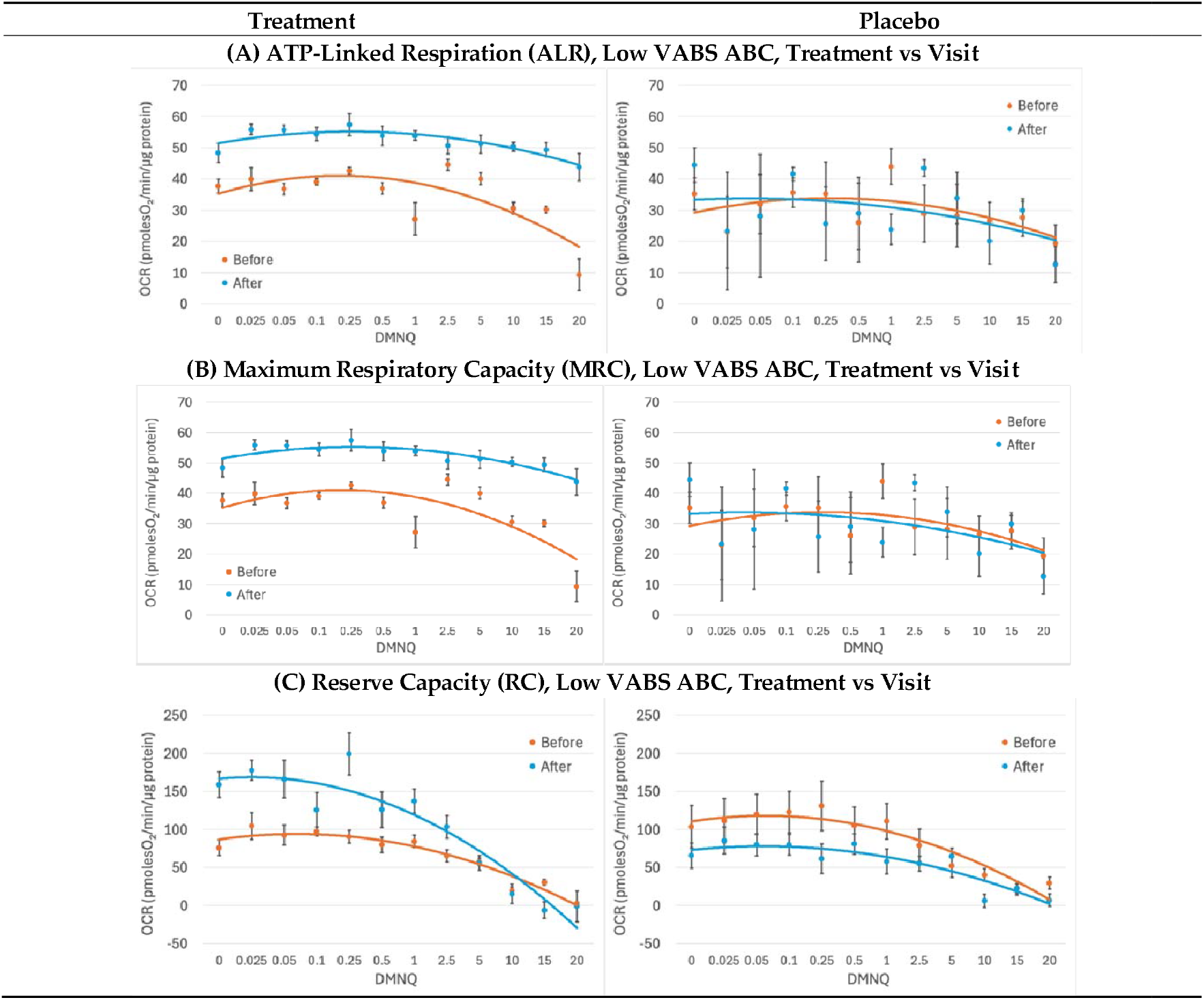
Low VABS ABC Mitochondria Effects. Effects were measured over the course of 12 weeks before and after treatment and placebo. Standard error bars are standard error of the mean. Mixed-model regression was used to control for the repeated subjects level effects while determining whether the effect of treatment vs placebo affected the change over time. Oxygen consumption rate (OCR); µM 2,3-dimethoxy-1,4-napthoquinone (DMNQ); Vineland Adaptive Behavior Scale (VABS).

ALR increased independent of physiological stress with the mitochondrial-targeted supplement, but not with placebo, but only for the low-functioning subjects [treatment x visit x VABC (high vs low): ALR F(1,1326)=29.35,p<0.001] (Figure 4A). No effect on ALR was noted in high-functioning subjects (Supplemental Figure 2). Thus, the mitochondrial-targeted supplement improved the ability of the mitochondria to produce ATP for those that were low functioning.

The supplement affected the average MRC and RC [treatment x visit x VABC (high vs low): MRC: F(1,1332)=87.55,p<0.001; RC: F(1,1332)=48.30,p<0.001] and the change in MRC and RC with increasing physiological stress [treatment x visit x DMNQ x VABC (high vs low): MRC: F(1,1323)=13.87, p<0.001; RC: F(1,1323)=20.39, p<0.001] differently for those that were low versus high functioning. Treatment increased the average MRC and RC and allowed the RMC and RC to maintain high respiration as physiological stress increased in low functioning individuals (Figure 4B-C), but not in high functioning individuals (Supplemental Figure 2).

In low functioning individuals, the MRC after treatment was maintained above the MRC before treatment regardless of the level of physiological stress. However, for low functioning individuals, RC after the treatment dipped below the RC before treatment for high physiological stress levels. This suggests that the mitochondrial-targeted supplements improved mitochondrial resistance to mild and moderate physiological stress in low-functioning individuals with ASD.

### 3.4. Behavioral and Cognitive Outcomes

The statistical results of behavioral and cognitive outcomes measures are provided in Table 2.

**Table 2.**
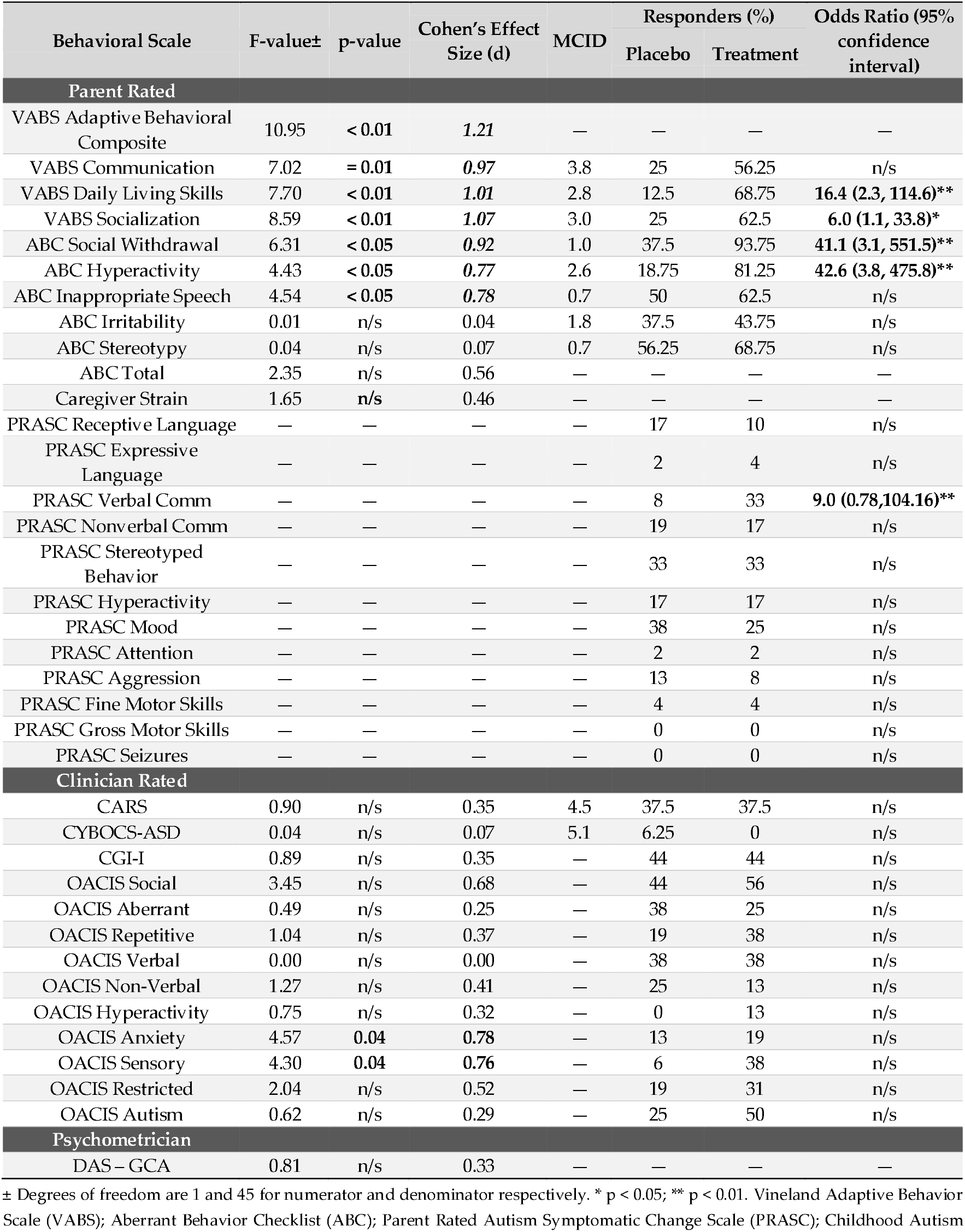
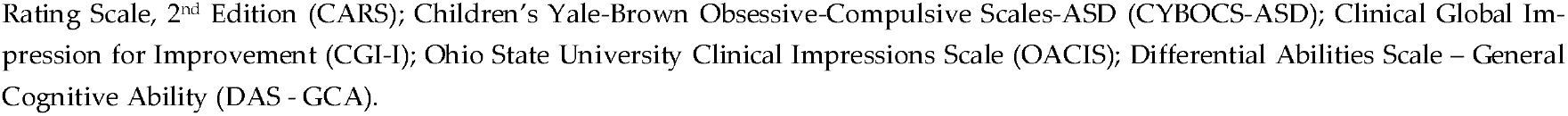
Statistics of Outcome Measures. Dashes indicate that no statistical test was performed.

| Behavioral Scale | F-value± | p-value | Cohen's Effect Size (d) | MCID | Responders (%) |  | Odds Ratio (95% confidence interval) |
| --- | --- | --- | --- | --- | --- | --- | --- |
|  |  |  |  |  | Placebo | Treatment |  |
| Parent Rated |  |  |  |  |  |  |  |
| VABS Adaptive Behavioral Composite | 10.95 | < 0.01 | 1.21 | — | — | — | — |
| VABS Communication | 7.02 | = 0.01 | 0.97 | 3.8 | 25 | 56.25 | n/s |
| VABS Daily Living Skills | 7.70 | < 0.01 | 1.01 | 2.8 | 12.5 | 68.75 | 16.4 (2.3, 114.6)** |
| VABS Socialization | 8.59 | < 0.01 | 1.07 | 3.0 | 25 | 62.5 | 6.0 (1.1, 33.8)* |
| ABC Social Withdrawal | 6.31 | < 0.05 | 0.92 | 1.0 | 37.5 | 93.75 | 41.1 (3.1, 551.5)** |
| ABC Hyperactivity | 4.43 | < 0.05 | 0.77 | 2.6 | 18.75 | 81.25 | 42.6 (3.8, 475.8)** |
| ABC Inappropriate Speech | 4.54 | < 0.05 | 0.78 | 0.7 | 50 | 62.5 | n/s |
| ABC Irritability | 0.01 | n/s | 0.04 | 1.8 | 37.5 | 43.75 | n/s |
| ABC Stereotypy | 0.04 | n/s | 0.07 | 0.7 | 56.25 | 68.75 | n/s |
| ABC Total | 2.35 | n/s | 0.56 | — | — | — | — |
| Caregiver Strain | 1.65 | n/s | 0.46 | — | — | — | — |
| PRASC Receptive Language | — | — | — | — | 17 | 10 | n/s |
| PRASC Expressive Language | — | — | — | — | 2 | 4 | n/s |
| PRASC Verbal Comm | — | — | — | — | 8 | 33 | 9.0 (0.78,104.16)** |
| PRASC Nonverbal Comm | — | — | — | — | 19 | 17 | n/s |
| PRASC Stereotyped Behavior | — | — | — | — | 33 | 33 | n/s |
| PRASC Hyperactivity | — | — | — | — | 17 | 17 | n/s |
| PRASC Mood | — | — | — | — | 38 | 25 | n/s |
| PRASC Attention | — | — | — | — | 2 | 2 | n/s |
| PRASC Aggression | — | — | — | — | 13 | 8 | n/s |
| PRASC Fine Motor Skills | — | — | — | — | 4 | 4 | n/s |
| PRASC Gross Motor Skills | — | — | — | — | 0 | 0 | n/s |
| PRASC Seizures | — | — | — | — | 0 | 0 | n/s |
| Clinician Rated |  |  |  |  |  |  |  |
| CARS | 0.90 | n/s | 0.35 | 4.5 | 37.5 | 37.5 | n/s |
| CYBOCS-ASD | 0.04 | n/s | 0.07 | 5.1 | 6.25 | 0 | n/s |
| CGI-I | 0.89 | n/s | 0.35 | — | 44 | 44 | n/s |
| OACIS Social | 3.45 | n/s | 0.68 | — | 44 | 56 | n/s |
| OACIS Aberrant | 0.49 | n/s | 0.25 | — | 38 | 25 | n/s |
| OACIS Repetitive | 1.04 | n/s | 0.37 | — | 19 | 38 | n/s |
| OACIS Verbal | 0.00 | n/s | 0.00 | — | 38 | 38 | n/s |
| OACIS Non-Verbal | 1.27 | n/s | 0.41 | — | 25 | 13 | n/s |
| OACIS Hyperactivity | 0.75 | n/s | 0.32 | — | 0 | 13 | n/s |
| OACIS Anxiety | 4.57 | 0.04 | 0.78 | — | 13 | 19 | n/s |
| OACIS Sensory | 4.30 | 0.04 | 0.76 | — | 6 | 38 | n/s |
| OACIS Restricted | 2.04 | n/s | 0.52 | — | 19 | 31 | n/s |
| OACIS Autism | 0.62 | n/s | 0.29 | — | 25 | 50 | n/s |
| Psychometrician |  |  |  |  |  |  |  |
| DAS – GCA | 0.81 | n/s | 0.33 | — | — | — | — |

± Degrees of freedom are 1 and 45 for numerator and denominator respectively. \* $p < 0.05$ ; \*\* $p < 0.01$ . Vineland Adaptive Behavior Scale (VABS); Aberrant Behavior Checklist (ABC); Parent Rated Autism Symptomatic Change Scale (PRASC); Childhood Autism
Rating Scale, 2<sup>nd</sup> Edition (CARS); Children’s Yale-Brown Obsessive-Compulsive Scales-ASD (CYBOCS-ASD); Clinical Global Impression for Improvement (CGI-I); Ohio State University Clinical Impressions Scale (OACIS); Differential Abilities Scale – General Cognitive Ability (DAS - GCA).

The VABS VABC, communication, daily living skills and socialization (Figure 5A-D) significantly improved with very-large effect sizes with the supplement as compared to the placebo (Table 2).

**Figure 5.**
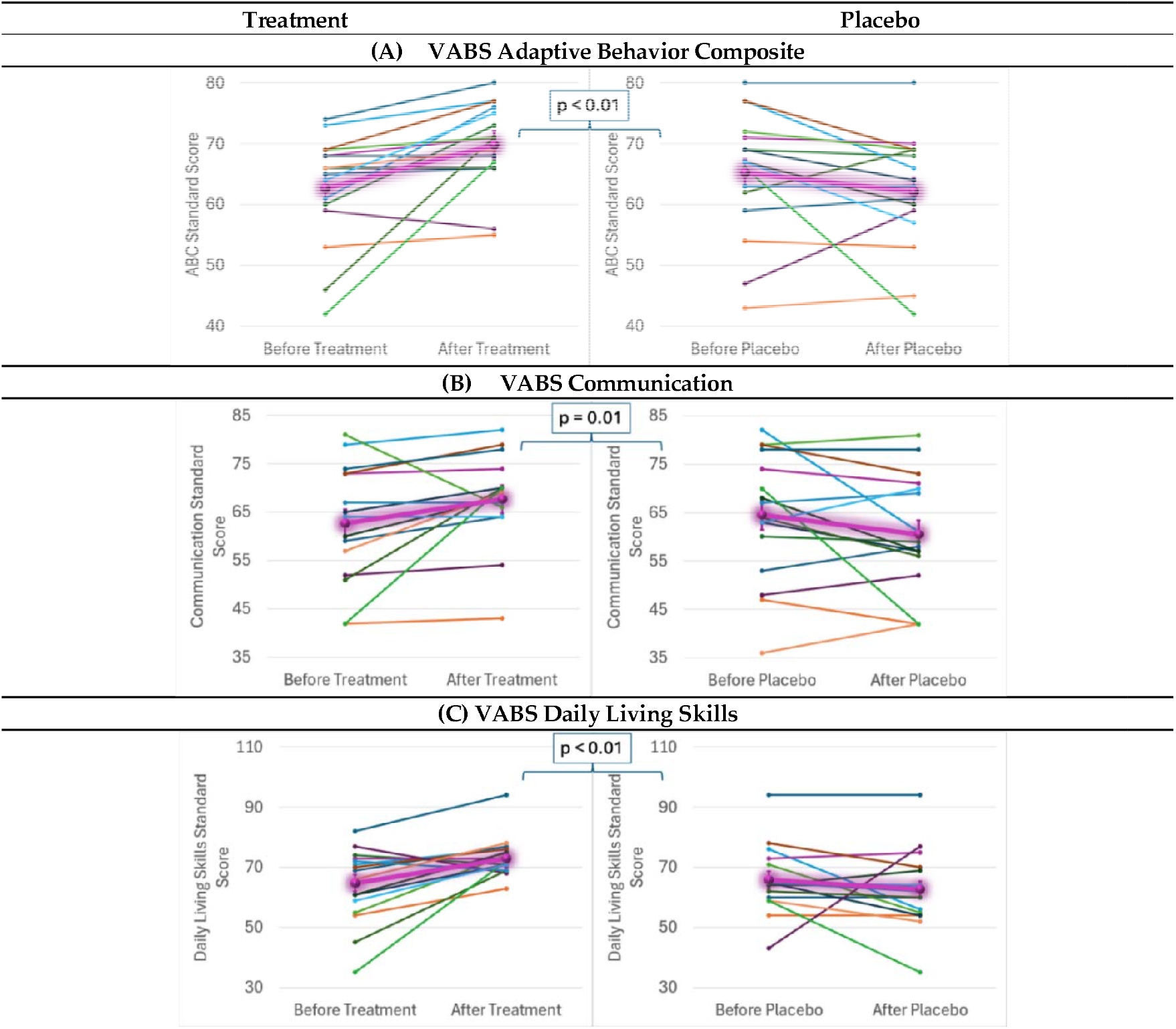
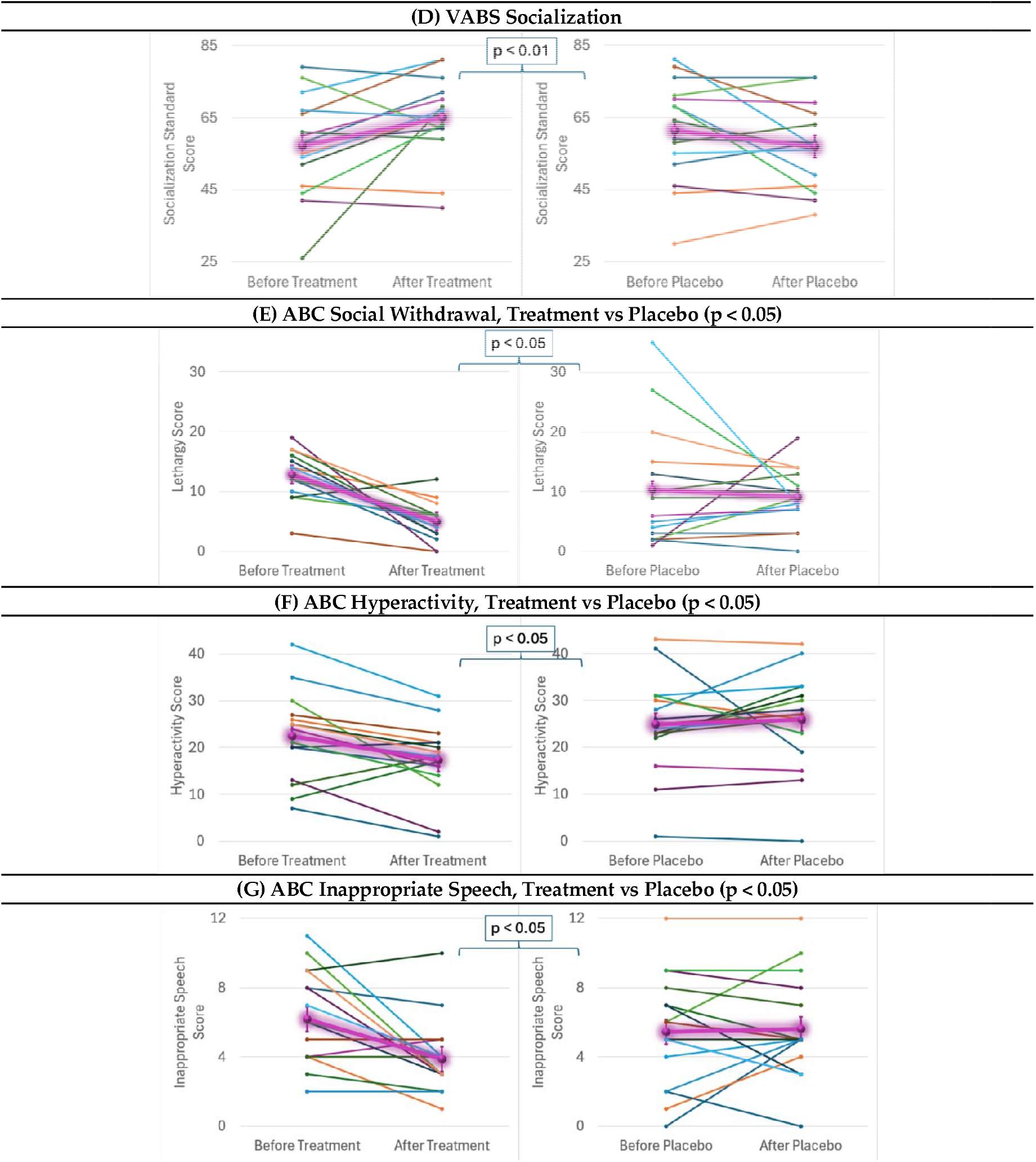
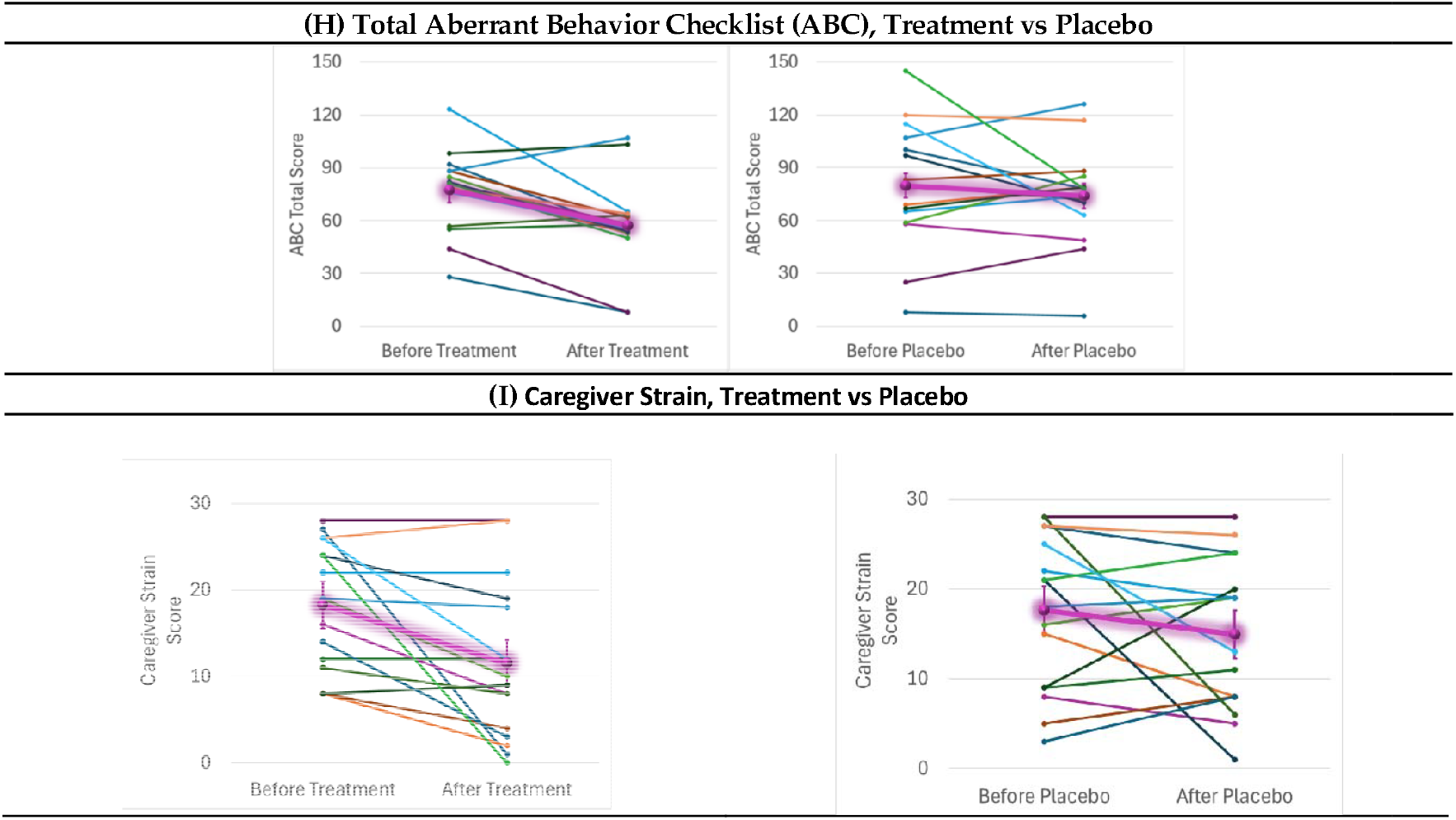
Behavior Scales Effects. Effects were measured over the course of 12 weeks before and after treatment and placebo. The highlighted purple lines represent the average effect for the group. Standard error bars are standard error of the mean. Mixed-model regression was used to control for the repeated subjects level effects while determining whether the effect of treatment vs placebo affected the change over time. Vineland Adaptive Behavior Scale (VABS); Aberrant Behavior Checklist (ABC).

The supplement improved ABC social withdrawal, hyperactivity and inappropriate speech (Figure 5E-G) with large effect sizes. Irritability and stereotypy (Supplement Figure 2) and total ABC (Figure 5H) scores improved with the supplement but were not statistically significant. Total ABC, which was borderline significant, had a medium effect size.

The supplement did not significantly reduce caregiver strain (Figure 5I) as compared to the placebo, although a trend for improvement was noted. The supplement did not significantly affect the PRAS-ASD or the SP2 although both improved more with the supplements (Supplementary Figure 4,5).

Two of the OACIS severity scales, anxiety [placebo before: 3.0 (1.6), after 3.4 (1.3), supplement before 3.5 (1.4), after 2.8 (1.5)] and sensory [placebo before: 4.3 (1.4), after 4.8 (1.7), supplement before 4.3 (1.7), after 3.6 (2.0)], improved with the supplement with medium-to-large effect sizes. Neither the CARS (Supplement Figure 6) nor the GCA (Supplement Figure 7) changed with the supplement as compared to placebo. The supplement had little effect on the CYBOS-ASD (Supplement Figure 8).

### 3.5. Responder analysis

More children receiving the supplement had a clinically-significant response on the VABS daily living skills and socialization subscales, ABC social withdrawal and hyper-activity subscales and PRASC verbal communication subscale as compared to the placebo (Table 2). There was no difference in responders for the clinician and psychometrician-rated instruments.

### 3.6. Adverse Events

One patient had mild transient constipation, diarrhea, and acid reflux on the mitochondrial supplement. There were no serious adverse events. There were no suicidal attempts or ideations, or significant self-injurious behaviors. The MDOTES found that participants reported decreased appetite significantly more often when taking the supplement. As these were not new events but rather mild worsening of ongoing symptoms, they were not considered AEs (See Supplementary Tables 2,3,4).

## 4. Discussion

Both mitochondrial disease and mitochondrial dysfunction have been linked to ASD.^5^ Children with ASD are not uncommonly treated for these abnormalities with various supplements but data supporting this practice is limited. This is the first controlled, blinded study to demonstrate that a safe and well-tolerated mitochondrial-targeted supplement can improve both mitochondrial function and symptoms in children with non-syndromic ASD. The supplement was found to partially normalize mitochondrial enzymology and to improve mitochondrial resilience, especially in lower-functioning children. Improvements in socialization, activities of daily living and hyperactivity exceeded the clinically important difference, demonstrating that these clinical changes were not only statistically significant but also clinically important.

### 4.1. Targeted Supplementation Improves Mitochondrial Function

The primary aim of this study was to determine whether the supplement improved mitochondrial function, as measured by buccal cell enzymology and respiration in PBMCs.

Before treatment, CS was markedly elevated, and ETC complex I, II and IV were markedly depressed. After receiving the supplement, CS partially normalized. The elevated CS indicates mitochondrial proliferation which presumably occurs to compensate for poorly functioning mitochondria. Partial normalization of CS indicates that the mitochondrial-targeted supplementation was able to partially restore normal mitochondrial health with 12 weeks of treatment. However, the ETC activity corrected for CS did not change much suggesting that the intrinsic deficits in mitochondrial physiology were not repaired but rather the mitochondria were better able to function with these deficits. Such an effect may have occurred by supporting reactive oxygen species control system and/or supported mitochondrial repair mechanism including fission, fusion and mitophagy to produce more optimal mitochondria. Regardless of the mechanism, these data suggest that mitochondrial function can be partially changed in a short period in children with ASD. Longer treatment may have had a greater effect on normalizing mitochondrial CS and may lead to changes in ETC activity also.

In this study, we found that a mitochondria-targeted supplement increased PBMC respiratory rates, with some respiratory parameters becoming more resilient to physiological stress. For example, as seen in Figure 4, ALR remained steady with increased physiological stress with supplement treatment, while ALR decreased with physiological stress with placebo treatment. With supplement treatment, PLR increased with physiological stress. This most likely indicates the recruitment of proton leak to control physiological stress at the inner mitochondrial membrane. Interestingly, variability increased in ALR and PLR after the placebo. We hypothesize that the standard errors in the placebo curves are larger because there was no treatment to stabilize the mitochondrial function, resulting in a more variable outcome. With supplement treatment, MRC and RC remained higher than the baseline MRC and RC throughout the physiological stress challenge with the exception that RC did lose this elevation at the highest level of physiological stress. When the participants were separated by developmental ability using the VABC, we found that these observed effects were driven by the lower-functioning individuals with ASD.

The MOST was developed to examine the response of the mitochondria to physiological stress.^5,8^ This assay allowed the discovery that LCLs from a subset of about 33% of children with ASD (called AD-A LCLs), have elevated respiratory rates including double that usual RC.^5,8^ Using the MOST, we previously found greater sensitivity to physiological stress when respiratory rates were elevated. PBMCs derived from a subset of children with ASD and neurodevelopmental regression were also found to have this characteristic.^7^ Interestingly, this same pattern of elevated respiratory rates and greater vulnerability can be induced with prolonged exposure to low levels of oxidative stress in LCLs^51^, and are associated with prenatal exposure to air pollution^42^ or lack of nutritional metal prenatally^40^ in PBMCs from ASD patients.

### 4.2. Targeted Mitochondrial Supplementation Improves Core ASD Symptoms

Several parent-completed outcomes were significant with large effects, including communication, socialization and daily living skills as rated with the VABS, and social withdrawal and hyperactivity as rated with the ABC. All these changes exceeded the MCID, expect communication, indicating that they were both statistically and clinically significant. Although communication did not surpass the MCID, it was found to be rated as improved at a significantly higher rate on the PRASC during the active treatment than during placebo.

Through observation and parental questioning, clinicians judged that the severity of anxiety and sensory issues reduced with the supplement treatment, but they did not note improvements in other areas that were found to be improved on the parent’s rated instruments. It should be appreciated that the clinician only observed the child for a short period of time in an unfamiliar environment, so the changes might not be obvious in this artificial environment. Testing of general cognitive abilities did not improve on the DAS. Such tests require adequate attention and engagement, which may have been subpar.

### 4.3. Comparing to other Treatment Studies

Some of the mainstay treatments for mitochondrial disease are used to treat children with ASD. Two DPBC studies using higher carnitine doses (N=30, 100mg/kg/d,^10^; N=30, 50mg/k/d^9^) than the current study (10-25mg/kg/d) found improvement in the CARS. Two DBPC studies which added carnitine to risperidone, found improvements in ABC sub-scales (hyperactivity, social withdrawal, inappropriate speech) similar to the current study.^11,12^

An open-label study using 400mg/kg/day of carnitine^13^ and another using 100mg/d of ubiquinol^15^ found improvements in verbal communication, similar to the current study, while another study found that improvement in oxidative stress as the results of treating with ubiquinone 30-60mg/d was associated with an improvement in CARS scores.^16^

A controlled study which examined a vitamin/mineral supplement similar to the treatment used in this study demonstrated parental-reported improvements in hyperactivity, tantruming and receptive language as well as improvements in ATP, NADH and NADPH blood concentrations.^52^

Only one study treated selected individuals with ASD and mitochondrial dysfunction using a method similar to the current study. The treatment contained carnitine, coenzyme Q10 and alpha-lipoic acid.^18^ Similar to the current study, improvements occurred in social withdrawal and inappropriate speech subscales of the ABC as well as mitochondrial activity.

In general, the current study expands on these previous studies to confirm and further extend the evidence for a mitochondrial targeted supplement improving core and associated symptoms of ASD. However, we note that previous studies were very variable in the treatment used and methodology, so a direct comparison is not possible. This is the first blinded study to show that a supplement can have direct improvement in mitochondrial function and ASD symptoms.

### 4.4. The Importance of Treating Mitochondrial Abnormalities in Neurodevelopmental Disorders

Mitochondrial dysfunction is highly prevalent in ASD with estimates ranging from about 30%^5,6^ to 80%.^53^ Although this study was limiting to those with abnormal mitochondrial activity, it does not limit the applicability of these findings to a small subset of ASD with mitochondrial dysfunction. Indeed, almost 90% of the ASD population screened for this study were eligible due to the presence of mitochondrial dysfunction as defined by our inclusion criteria. One bias is that participants were managed by a child neurologist well-known in the ASD community for difficult, refractory cases, so the participants would tend to include more lower functioning individuals.

During prenatal neurodevelopment, mitochondria are central for all stages of neurogenesis, regulating neuronal length and complexity, synaptic function and hyperexcitability and have been linked to white matter connectivity.^5^ During postnatal development, the central nervous system heavily relies on multiple mitochondrially-mediated mechanisms, such as synaptic transmission, the glutamate-glutamine cycle and neurotransmitter homeostasis, the glutamate-GABA cycle, neuron-astrocyte shuttling of metabolic substrates like fatty acids and lactate, intercellular mitochondrial trafficking and mitophagy.^5^

### 4.5. Limitations

This study had limitations. First, the small sample size may have limited the sensitivity of the analyses to detect some treatment effects. Second, there was no washout period between the two arms of the study, so carryover effects could have occurred. Third, the single-site design only provides limited generalization of these results. Fourth, although no significant AEs were identified, tolerability data of the mitochondrial-targeted supplement combination would benefit from further study. Fifth, further studies will be needed to determine the optimal dose and treatment period.

## 5. Conclusions

Only two drugs are approved by the United States Food and Drug Administration for the treatment of ASD, both antipsychotic drugs which treat symptoms, not core biological abnormalities, and are associated with relatively common, serious short- and long-term AEs.^3^ Thus, well-tolerated interventions that target pathophysiological processes and core symptoms associated with ASD are sorely needed. As ASD is likely a lifelong disorder, the long-term adverse effects of any treatment are of concern.

In this small trial of children with non-syndromic ASD and mitochondrial dysfunction, treatment with a well-tolerated, dietary supplement combination designed to target mitochondrial function, for 12-weeks, resulted in improvement in mitochondrial function and parent-rated developmental and behavioral indexes. The areas of significant clinical improvement closely overlap with those in a previous study with another dietary supplement combination that also, in part, targeted mitochondria,^18,52^ thus supporting the validity of the present results. These promising findings should be considered preliminary until treatment is assessed in larger multicenter studies with longer duration.

## Data Availability

All data produced in the present study are available upon reasonable request to the authors.

## Acknowledgments

This research was supported by The Brain Foundation (Pleasanton, CA) and the EXL Foundation (Phoenix AZ). NeuroNeeds^®^ provided both active supplements and placebos only. None of the sponsors were involved with the design or conduct of the study, collection, management, analysis or interpretation of the data; or preparation, approval of the manuscript or decision to submit the manuscript for publication. Dr. Richard Boles is an owner and officer of NeuroNeeds^®^, as well as a clinician in private practice. He served as an advisor during study design, interpretation and publication, but had no role in human subjects or study procedures. All final decisions were made solely by Dr. Richard Frye from conception to submission. Dr. Frye is a member of the Scientific Advisory Board to The Brain Foundation. All other authors have no conflicts of interest to declare. The authors would like to thank all the staff which were involved in making this study possible and the participants and their families who gave their time.

## Author Contributions

Conceptualization, R.E.F., R.G.B.; data curation, P.J.M., Z.R.H.; analysis, R.E.F., P.J.M., Z.R.H.; funding acquisition, R.E.F., R.G.B.; investigation, R.E.F., R.G.B., P.J.M., Z.R.H.; methodology, R.E.F., R.G.B., P.J.M.; administration, R.E.F.; resources, R.E.F., R.G.B.; supervision, R.E.F.; validation, R.E.F., P.J.M.; visualization, Z.R.H.; writing – original draft, Z.R.H., R.E.F.; writing – review/edit, R.E.F., R.G.B., P.J.M., Z.R.H. All authors have read and agreed to the publication of this manuscript.

## Institutional Review Board Statement

This trial is registered on clinicaltrials.gov as NCT03835117 and performed under IND 142751.

## Informed Consent Statement

Parents of participants provided written informed consent.

## Disclaimer/Publisher’s Note

The statements, opinions and data contained in all publications are solely those of the individual author(s) and contributor(s) and not of MDPI and/or the editor(s). MDPI and/or the editor(s) disclaim responsibility for any injury to people or property resulting from any ideas, methods, instructions or products referred to in the content.

