## Supplemental Materials for "A Mitochondrial Supplement Improves Function and Mitochondrial Activity in Autism: A Double-Blind Place-bo-Controlled Cross-Over Trial"

**SUPPLEMENTAL MATERIALS (Ver 1.2 Jan 20 2025)**

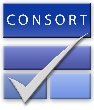
CONSORT 2010 checklist of information to include when reporting a randomised trial*

| Section/Topic | Item No | Checklist item | Reported on page No |
| --- | --- | --- | --- |
| Title and abstract | | | |
|  | 1a | Identification as a randomised trial in the title | 1 |
|  | 1b | Structured summary of trial design, methods, results, and conclusions (for specific guidance see CONSORT for abstracts) | 2 |
| Introduction | | | |
| Background and objectives | 2a | Scientific background and explanation of rationale | 3-5 |
|  | 2b | Specific objectives or hypotheses | 4-5 |
| Methods | | | |
| Trial design | 3a | Description of trial design (such as parallel, factorial) including allocation ratio | 5-6 |
|  | 3b | Important changes to methods after trial commencement (such as eligibility criteria), with reasons | 5 |
| Participants | 4a | Eligibility criteria for participants | 8-9 |
|  | 4b | Settings and locations where the data were collected | 5 |
| Interventions | 5 | The interventions for each group with sufficient details to allow replication, including how and when they were actually administered | 6-8 |
| Outcomes | 6a | Completely defined pre-specified primary and secondary outcome measures, including how and when they were assessed | 9-17 |
|  | 6b | Any changes to trial outcomes after the trial commenced, with reasons | 5 |
| Sample size | 7a | How sample size was determined | 9 |
|  | 7b | When applicable, explanation of any interim analyses and stopping guidelines | 13 |
| Randomisation: |  |  |  |
| Sequence generation | 8a | Method used to generate the random allocation sequence | 6 |
|  | 8b | Type of randomisation; details of any restriction (such as blocking and block size) | 6 |
| Allocation concealment mechanism | 9 | Mechanism used to implement the random allocation sequence (such as sequentially numbered containers), describing any steps taken to conceal the sequence until interventions were assigned | 6 |
| Implementation | 10 | Who generated the random allocation sequence, who enrolled participants, and who assigned participants to interventions | 6 |
| Blinding | 11a | If done, who was blinded after assignment to interventions (for example, participants, care providers, those assessing outcomes) and how | 6 |
|  | 11b | If relevant, description of the similarity of interventions | 8 |
| Statistical methods | 12a | Statistical methods used to compare groups for primary and secondary outcomes | 17-18 |
|  | 12b | Methods for additional analyses, such as subgroup analyses and adjusted analyses | 17-18 |
| Results | | | |
| Participant flow (a diagram is strongly recommended) | 13a | For each group, the numbers of participants who were randomly assigned, received intended treatment, and were analysed for the primary outcome | 19 |
|  | 13b | For each group, losses and exclusions after randomisation, together with reasons | 19 |
| Recruitment | 14a | Dates defining the periods of recruitment and follow-up | 6 |
|  | 14b | Why the trial ended or was stopped |  |
| Baseline data | 15 | A table showing baseline demographic and clinical characteristics for each group | 20-21 |
| Numbers analysed | 16 | For each group, number of participants (denominator) included in each analysis and whether the analysis was by original assigned groups | 20-21 |
| Outcomes and estimation | 17a | For each primary and secondary outcome, results for each group, and the estimated effect size and its precision (such as 95% confidence interval) | 16-26 |
|  | 17b | For binary outcomes, presentation of both absolute and relative effect sizes is recommended | 22,26 |
| Ancillary analyses | 18 | Results of any other analyses performed, including subgroup analyses and adjusted analyses, distinguishing pre-specified from exploratory | 22-26 |
| Harms | 19 | All important harms or unintended effects in each group (for specific guidance see CONSORT for harms) | 34-35 |
| Discussion | | | |
| Limitations | 20 | Trial limitations, addressing sources of potential bias, imprecision, and, if relevant, multiplicity of analyses | 41 |
| Generalisability | 21 | Generalisability (external validity, applicability) of the trial findings | 38-39 |
| Interpretation | 22 | Interpretation consistent with results, balancing benefits and harms, and considering other relevant evidence | 35-41 |
| Other information | | |  |
| Registration | 23 | Registration number and name of trial registry | 1 |
| Protocol | 24 | Where the full trial protocol can be accessed, if available | Supplement |
| Funding | 25 | Sources of funding and other support (such as supply of drugs), role of funders | 43 |

*We strongly recommend reading this statement in conjunction with the CONSORT 2010 Explanation and Elaboration for important clarifications on all the items. If relevant, we also recommend reading CONSORT extensions for cluster randomised trials, non-inferiority and equivalence trials, non-pharmacological treatments, herbal interventions, and pragmatic trials. Additional extensions are forthcoming: for those and for up to date references relevant to this checklist, see [www.consort-statement.org](http://www.consort-statement.org).

| 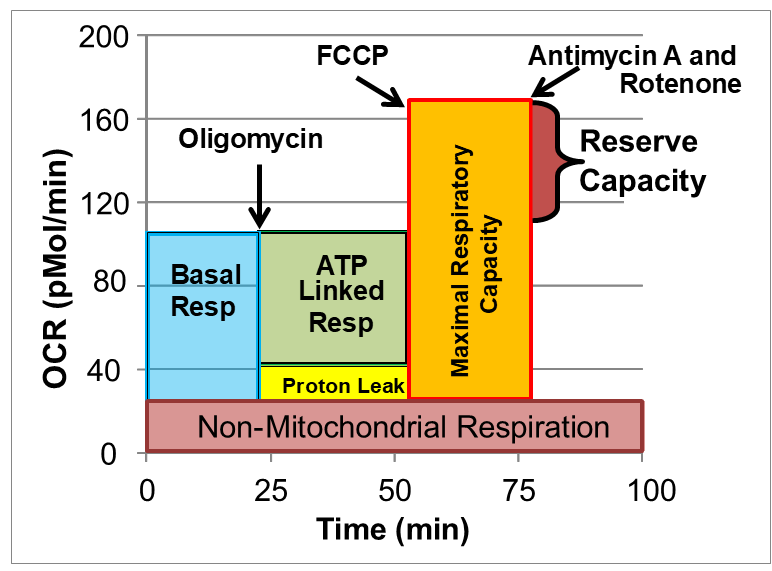 | **Supplementary Figure 1.** Seahorse assay and various mitochondrial respiratory parameters. Oxygen consumption rate (OCR) is measured to determine mitochondrial activity. Three OCRs are measured over 18 minutes to determine mitochondrial activity for each segment of the assay. Regents are added to determine parameters of mitochondrial activity. Basal Respiration is the difference between baseline OCR and non-mitochondrial OCR. Oligomycin, which is a complex V inhibitor, is added to determine the portion of Basal Respiration that is ATP-Linked Respiration (ALR) and Proton-Leak Respiration (PLR). Carbonyl cyanide-p-trifluoromethoxyphenyl-hydrazone (FCCP), a protonophore, is added to collapse the inner membrane gradient, driving the mitochondria to respire at its maximal rate. This determines Maximal Respiratory Capacity (MRC). Antimycin A and Rotenone, complex III and I inhibitors, stop mitochondrial respiration to determine the non-mitochondrial respiration. Reserve Capacity (RC) is the difference between Basal Respiration and Maximal Respiratory Capacity. Reprinted from. ^1^ |
| --- | --- |

**Supplemental Table 1. Demographic and Clinical Characteristics by Treatment Group.^1^ Mean values with standard deviation in parenthesis and range in brackets.**

| **Variable** | **Group A – Placebo then Treatment (n=9)** | **Group B – Treatment then Placebo**  **(n=7)** | **All Groups**  **(n=16)** |
| --- | --- | --- | --- |
| Age, mean (SD), years months | 9y 0m (3y 11m) | 9y 8m (4y 1m) | 9y 4m (5y 1m) |
| Males, N (%) | 7 (77.78%) | 7 (100%) | 14 (87.5%) |
| Percent Compliance, (%) |  |  |  |
| Placebo, SpectrumNeeds^®^ | 91.1% | 69.8% | 80.5% |
| Placebo, QNeeds^®^ | 96.3% | 89.3% | 92.8% |
| Treatment, SpectrumNeeds^®^ | 97.4% | 93.3% | 95.3% |
| Treatment, QNeeds^®^ | 94.7% | 96.4% | 95.5% |
| Race, N (%)^2^ |  |  |  |
| Asian | 1 (11.11%) | 1 (14.29%) | 2 (12.5%) |
| Black or African American | 0 (0%) | 0 (0%) | 0 (0%) |
| White | 5 (55.56%) | 4 (57.14%) | 9 (56.25%) |
| Other | 3 (33.33%) | 2 (28.57%) | 5 (31.25%) |
| Ethnicity, N (%) |  |  |  |
| Hispanic or Latino | 3 (33.33%) | 3 (42.86%) | 6 (37.5%) |
| Not Hispanic or Latino | 6 (66.67%) | 4 (57.14%) | 10 (62.5%) |
| Vineland Adaptive Behavior Composite, mean (SD) | 61.56 (11.58) | 64 (9.57) | 62.78 (15.02) |
| DAS GCA, mean (SD) | 62.11 (24.19) | 68.43 (27.96) | 65.27 (36.97) |
| Childhood Autism Rating Scale, mean (SD) | 38.11 (8.52) | 36.5 (3.75) | 37.31 (9.31) |
| Total Therapy, minutes per week, mean (SD) | 796.67 (1267.22) | 611.43 (1219.81) | 704.05 (1758.92) |
| Total Therapy, minutes per week, median [min; max] | 120 [0; 3480] | 120 [0; 3330] | 120 [0; 3480] |
| Speech Therapy, minutes per week, mean (SD) | 41.67 (33.35) | 40.71 (76.83) | 41.19 (83.76) |
| Behavioral Therapy, minutes per week, mean (SD) | 706.67 (1225.52) | 415.71 (937.30) | 561.19 (1542.86) |
| Physical Therapy, minutes per week, mean (SD) | 0 (0) | 21.43 (27.19) | 10.72 (27.19) |
| Occupational Therapy, minutes per week, mean (SD) | 28.33 (35.53) | 79.29 (127.49) | 53.81 (132.35) |
| Vision Therapy, minutes per week, mean (SD) | 3.33 (10) | 0 (0) | 1.67 (10) |
| Feeding Therapy, minutes per week, mean (SD) | 6.67 (20) | 0 (0) | 3.34 (20) |
| Social Group Therapy, minutes per week, mean (SD) | 3.33 (10) | 34.29 (90.71) | 18.81 (91.26) |
| Music Therapy, minutes per week, mean (SD) | 6.67 (20) | 7.14 (18.90) | 6.91 (27.52) |
| Citrate Synthase, mean (SD) | 34.13 (13.01) | 39.92 (3.75) | 37.03 (13.54) |
| Complex I, Uncorrected, mean (SD) | 135.31 (101.71) | 101.4 (63.27) | 118.36 (119.78) |
| Complex I, Corrected, mean (SD) | 3.61 (2.21) | 2.87 (1.88) | 3.24 (2.9) |
| Complex II, Uncorrected, mean (SD) | 1.56 (0.95) | 3.8 (0.6) | 2.68 (1.12) |
| Complex II, Corrected, mean (SD) | 0.05 (0.02) | 0.12 (0.07) | 0.09 (0.07) |
| Complex IV, Uncorrected, mean (SD) | 6.5 (2.84) | 6.88 (1.73) | 6.69 (3.33) |
| Complex IV, Corrected, mean (SD) | 0.2 (0.10) | 0.19 (0.05) | 0.20 (0.11) |
| Deficient Citrate Synthase, N (%) | 0 (0%) | 0 (0%) | 0 (0%) |
| Elevated Citrate Synthase, N (%) | 7 (77.78%) | 4 (57.14%) | 11 (68.75%) |
| Deficient Complex I, N (%) | 6 (66.67%) | 4 (57.14%) | 10 (62.5%) |
| Elevated Complex I, N (%) | 0 (0%) | 0 (0%) | 0 (0%) |
| Deficient Complex II, N (%) | 1 (11.11%) | 0 (0%) | 1 (6.25%) |
| Elevated Complex II, N (%) | 0 (0%) | 0 (0%) | 0 (0%) |
| Deficient Complex IV, N (%) | 2 (22.22%) | 0 (0%) | 2 (12.5%) |
| Elevated Complex IV, N (%) | 0 (0%) | 0 (0%) | 0 (0%) |
| Mitochondrial Disease, N (%) | 1 (11.11%) | 0 (0%) | 1 (6.25%) |
| Diagnostic Documentation, N (%) |  |  |  |
| Regression | 4 (44.44%) | 4 (57.14%) | 8 (50%) |
| History of Seizures | 0 (0%) | 1 (14.29%) | 1 (6.25%) |
| Mental Health Conditions | 1 (11.11%) | 2 (28.57%) | 3 (18.75%) |
| Serious Behavioral Problems | 3 (33.33%) | 1 (14.29%) | 4 (25%) |
| Allergies | 3 (33.33%) | 2 (28.57%) | 5 (31.25%) |
| Other Physical Conditions | 0 (0%) | 2 (28.57%) | 2 (12.5%) |
| Medications (Concurrent Treatments), N (%) |  |  |  |
| Levocarnitine | 2 (22.22%) | 0 (0%) | 2 (12.5%) |
| Naltrexone | 0 (0%) | 1 (14.29%) | 1 (6.25%) |
| Melatonin | 1 (11.11%) | 1 (14.29%) | 2 (12.5%) |
| Digestive Enzyme | 2 (22.22%) | 0 (0%) | 2 (12.5%) |
| Atomoxetine | 1 (11.11%) | 1 (14.29%) | 2 (12.5%) |
| MiraLAX | 1 (11.11%) | 0 (0%) | 1 (6.25%) |
| Hydroxyzine | 1 (11.11%) | 1 (14.29%) | 2 (12.5%) |
| Propranolol | 2 (22.22%) | 0 (0%) | 2 (12.5%) |
| Guanfacine | 0 (0%) | 1 (14.29%) | 1 (6.25%) |
| EpiPen | 0 (0%) | 1 (14.29%) | 1 (6.25%) |
| Galantamine | 1 (11.11%) | 0 (0%) | 1 (6.25%) |
| Cyproheptadine | 0 (0%) | 1 (14.29%) | 1 (6.25%) |
| Namenda | 1 (11.11%) | 0 (0%) | 1 (6.25%) |
| Naproxen | 0 (0%) | 1 (14.29%) | 1 (6.25%) |
| Leucovorin | 1 (11.11%) | 0 (0%) | 1 (6.25%) |
| Supplements (Concurrent Treatments), N (%) |  |  |  |
| Multivitamin | 1 (11.11%) | 0 (0%) | 1 (6.25%) |
| Probiotics | 2 (22.22%) | 0 (0%) | 2 (12.5%) |
| Vitamin D3 | 1 (11.11%) | 0 (0%) | 1 (6.25%) |
| Omega-3 | 1 (11.11%) | 0 (0%) | 1 (6.25%) |

^1^Baseline outcome measures outlined in Table 2 were not significantly different across treatment groups.

^2^No patients identifying as American Indian, Alaska Native, Hawaiian, or Pacific Islander were enrolled in the study.

| **Supplemental Figure 2: High VABS ABC Mitochondria Effects** | |
| --- | --- |
| Treatment | Placebo |
| **(A) ATP-Linked Respiration (ALR), High VABS ABC, Treatment vs Visit** | |
| 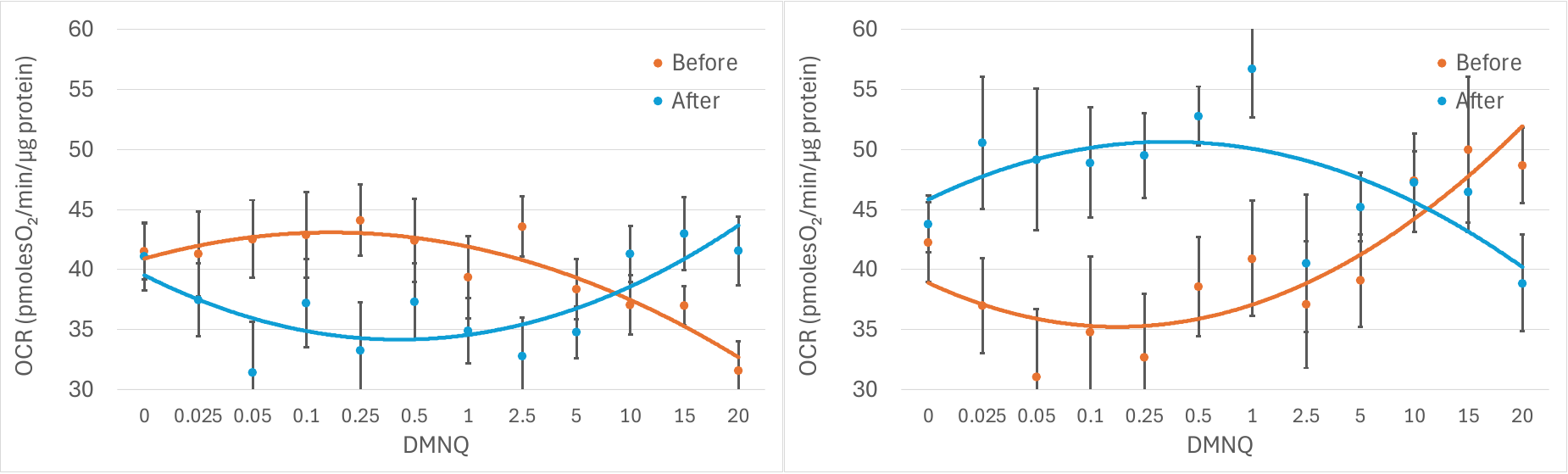 | |
| **(B) Maximum Respiratory Capacity (MRC), High VABS ABC, Treatment vs Visit** | |
| 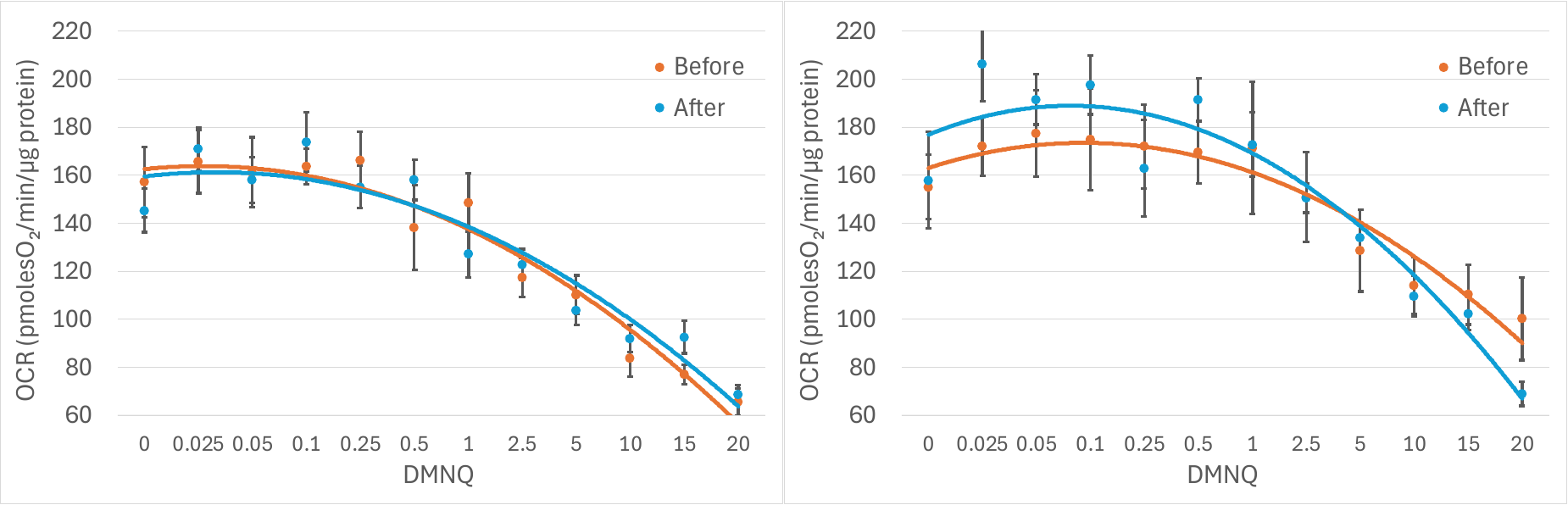 | |
| **(C) Reserve Capacity (RC), High VABS ABC, Treatment vs Visit** | |
| 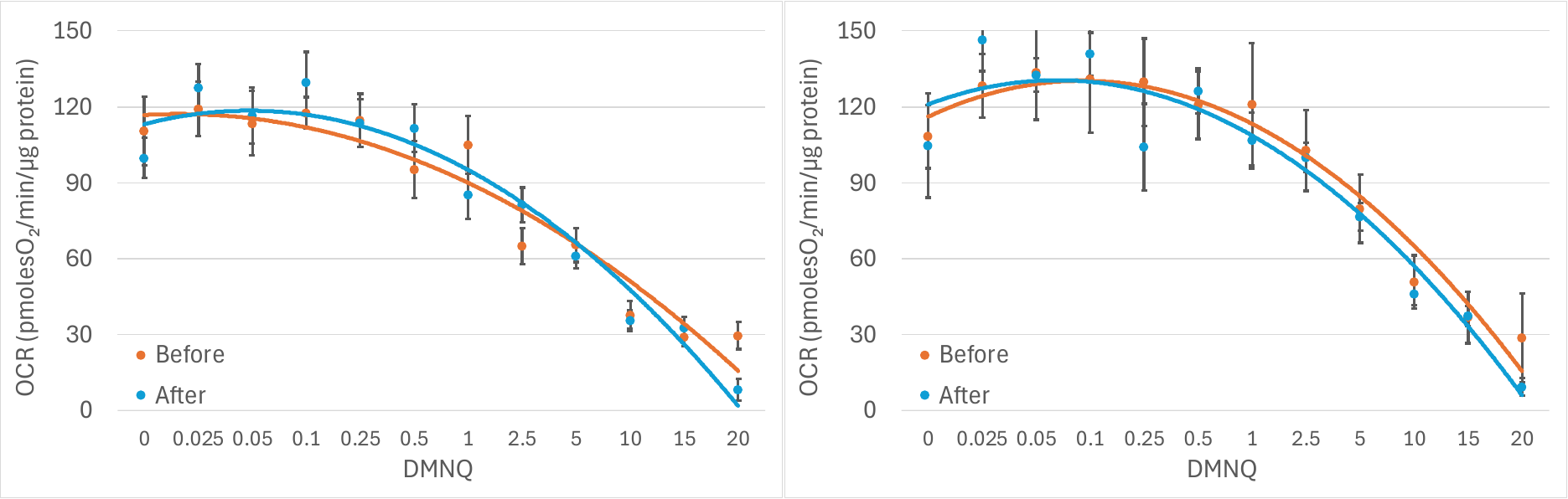 | |

| **Supplemental Figure 3: Behavior Scales**  The highlighted purple lines represent that average effect for the group | |
| --- | --- |
| Treatment | Placebo |
| 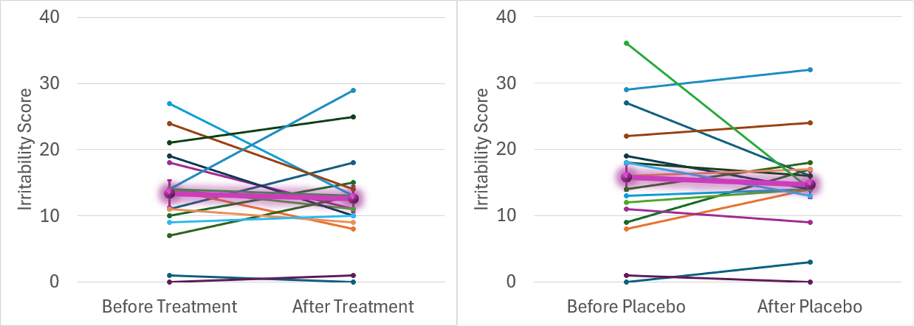 | |
| (A) **ABC Irritability, Treatment vs Placebo** | |
| 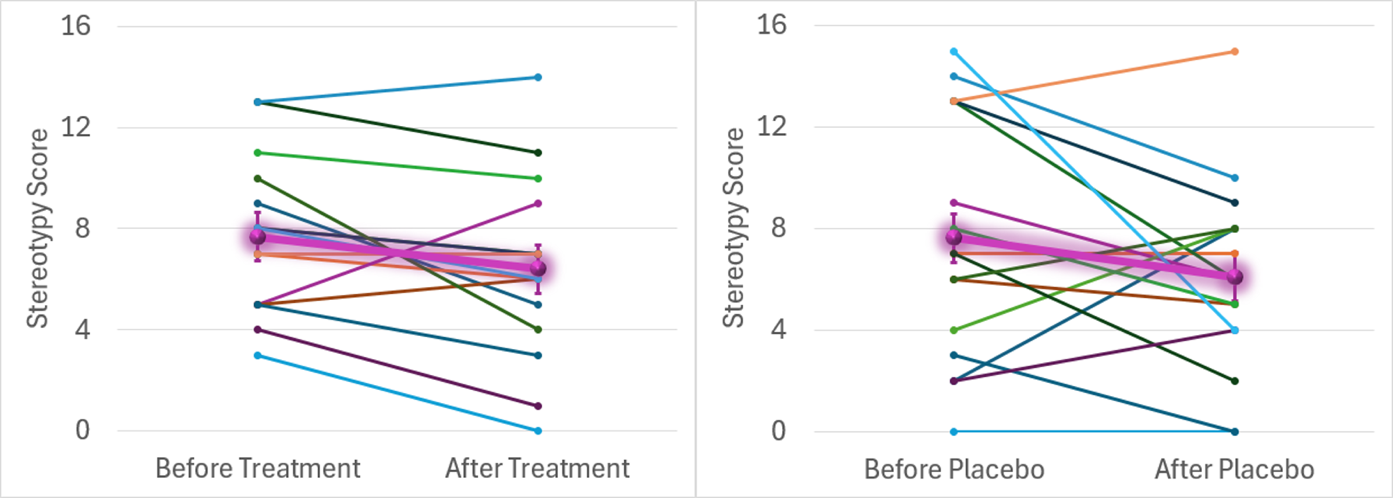 | |
| (B) **ABC Stereotypy, Treatment vs Placebo** | |

| Treatment | Placebo |
| --- | --- |
| 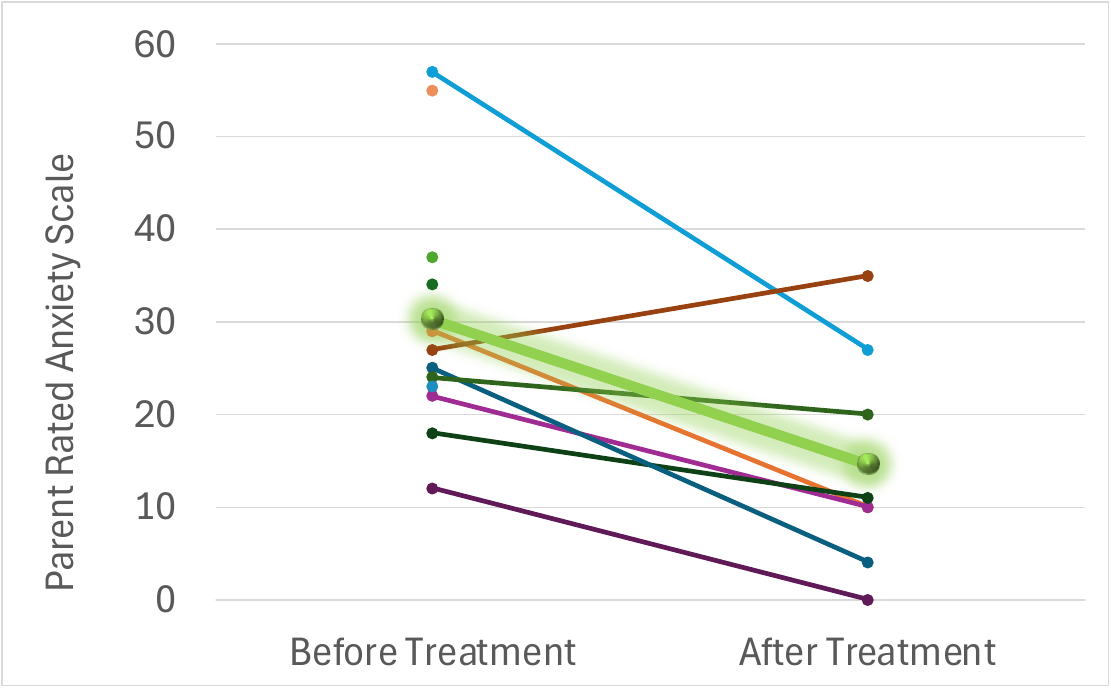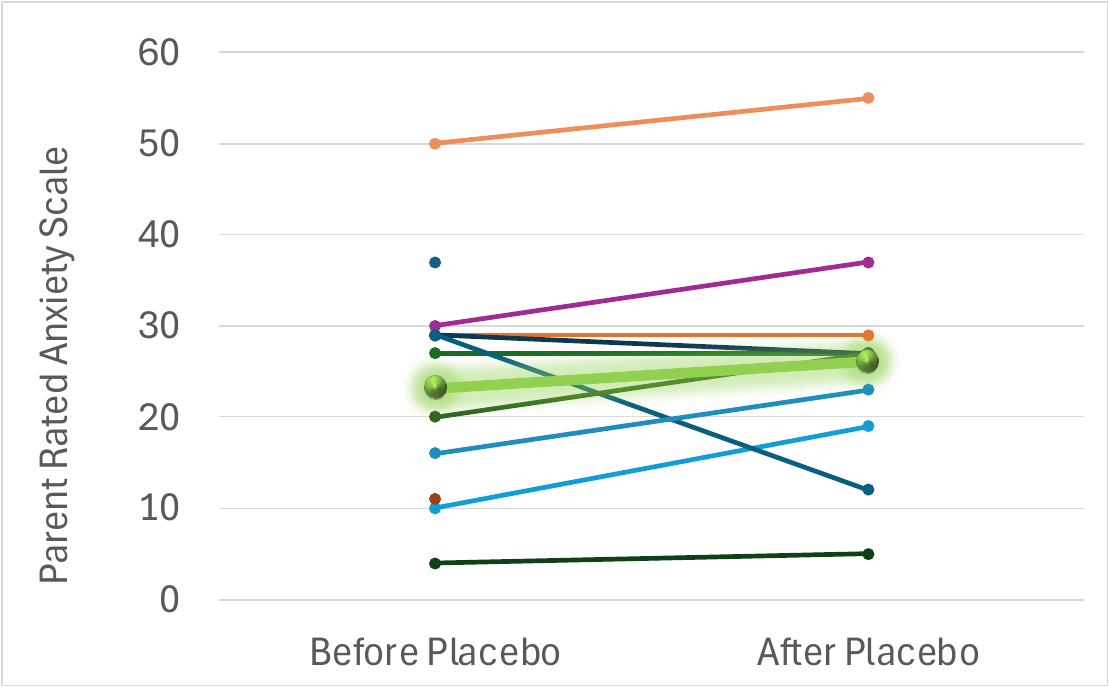 | |
| **Supplemental Figure 4: Parent-Rated Anxiety Scale (PRAS), Treatment vs Placebo** | |

| Treatment | Placebo |
| --- | --- |
| 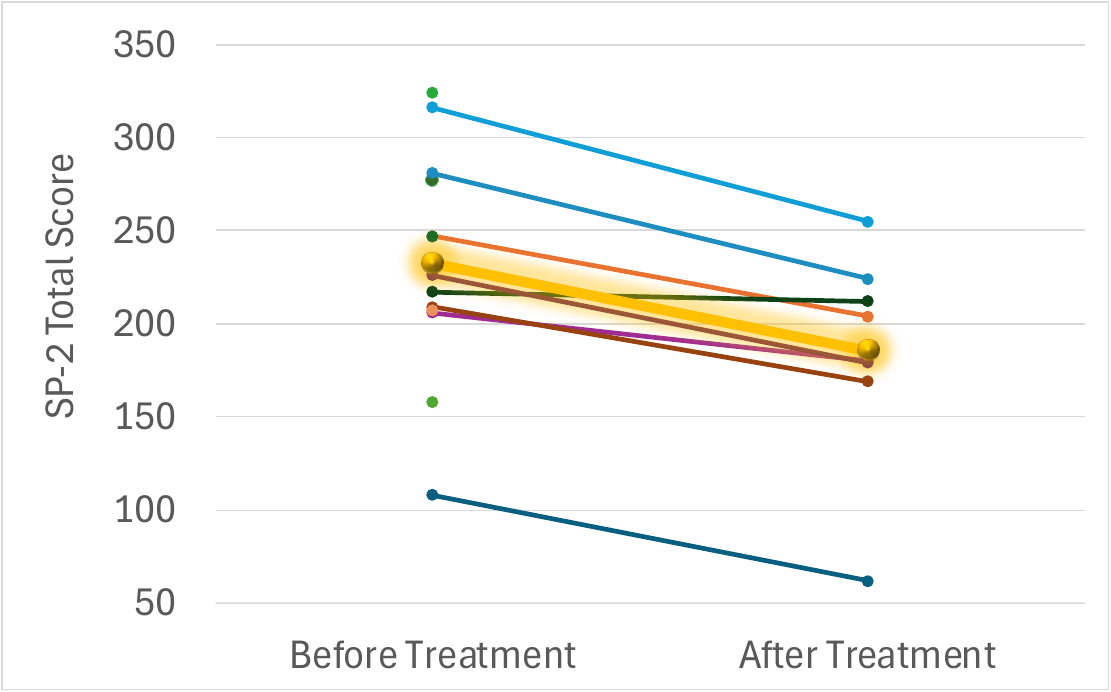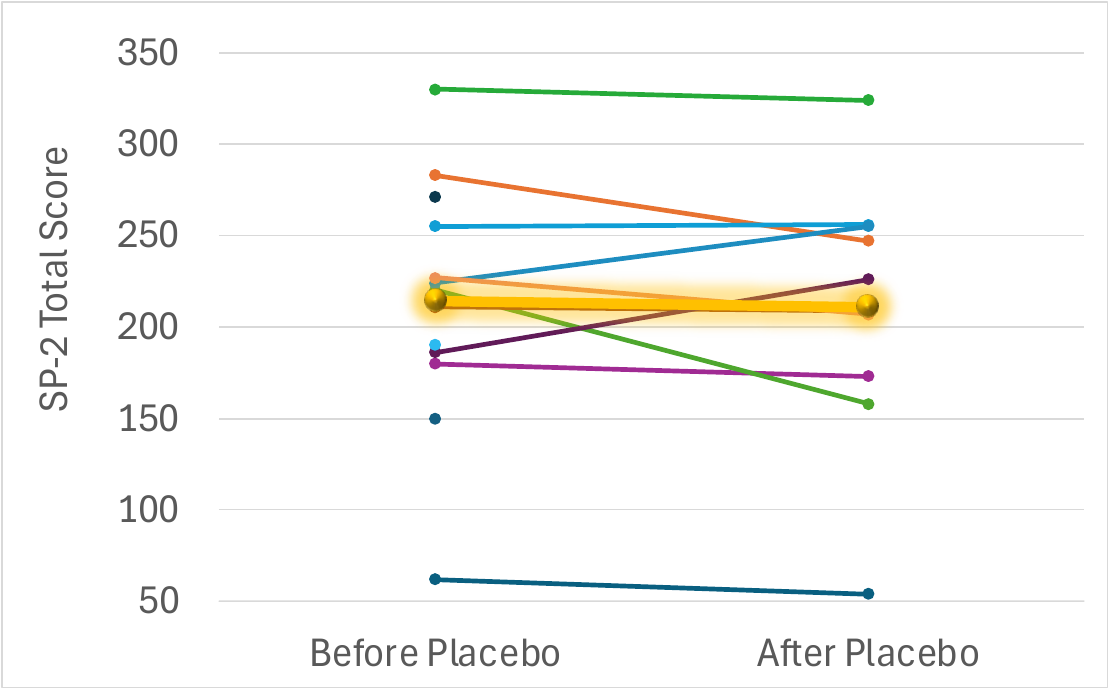 | |
| **Supplemental Figure 5: Sensory Profile 2 (SP-2), Treatment vs Placebo** | |

| Treatment | Placebo |
| --- | --- |
| 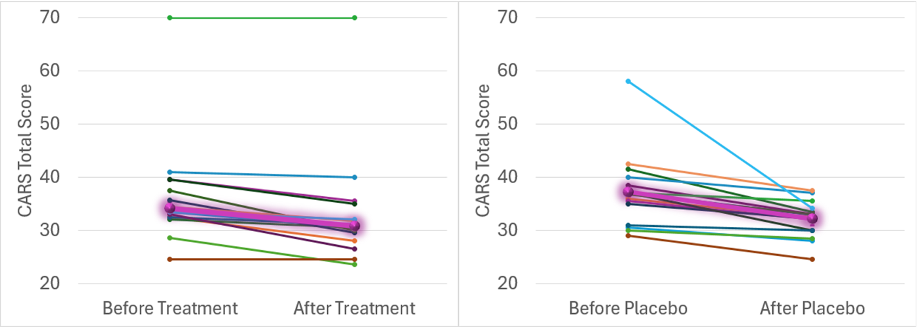 | |
| **Supplemental Figure 6: Childhood Autism Rating Scale (CARS), Treatment vs Placebo** | |

| Treatment | Placebo |
| --- | --- |
| 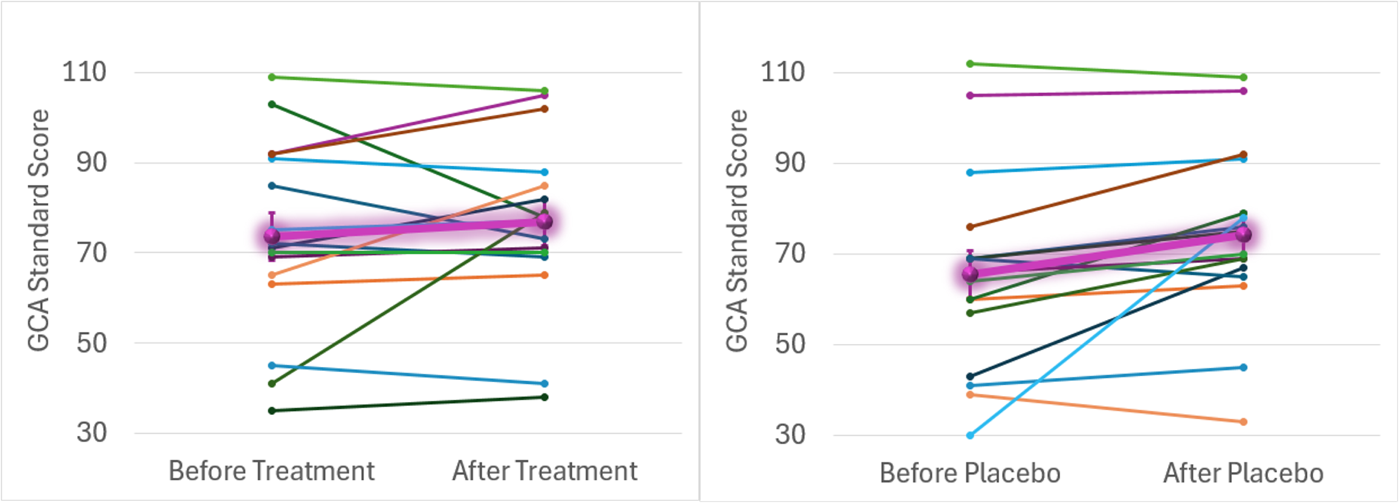 | |
| **Supplemental Figure 7: DAS General Cognitive Ability (GCA), Treatment vs Placebo** | |

| Treatment | Placebo |
| --- | --- |
| 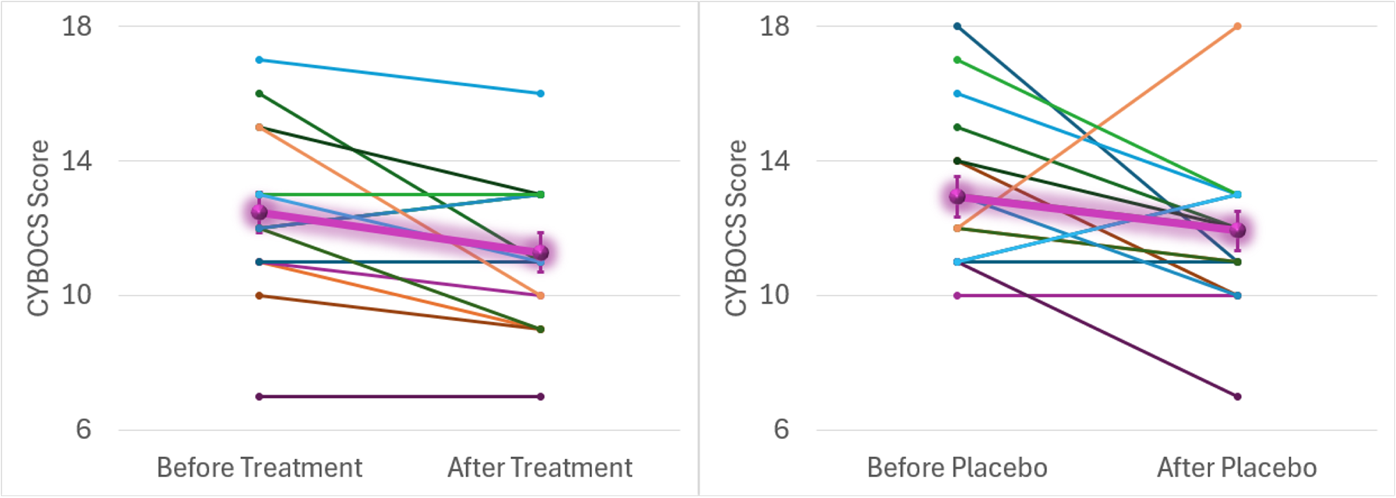 | |
| **Supplemental Figure 8: Children’s Yale-Brown Obsessive Compulsive Scale (CYBOCS), Treatment vs Placebo** | |

**Supplemental Table 2. Incidence of Symptoms Elicited by the modified Dosage Record Treatment Emergent Symptom (MDOTES) scale during the supplement and placebo treatment.**

| **Symptoms, N (%)** | **Supplement (n = 12)** | **Placebo (n = 13)** | **Overall (n = 25)** | **Fisher P** |
| --- | --- | --- | --- | --- |
| Any adverse effect | 11 (91.67) | 10 (76.92) | 21 (84) | 0.59 |
| Excitement/agitation | 7 (58.33) | 7 (53.85) | 14 (56) | 1.00 |
| Increased tantrums | 7 (58.33) | 7 (53.85) | 14 (56) | 1.00 |
| Emotional lability | 5 (41.67) | 4 (30.77) | 9 (36) | 0.69 |
| Increased motor activity | 3 (25) | 4 (30.77) | 7 (28) | 1.00 |
| Nasal congestion | 4 (33.33) | 3 (23.08) | 7 (28) | 1.00 |
| Aggression | 2 (16.67) | 5 (38.46) | 7 (28) | 0.32 |
| Insomnia | 1 (8.33) | 5 (38.46) | 6 (24) | 0.15 |
| Dry mouth/excessive thirst | 3 (25) | 3 (23.08) | 6 (24) | 1.00 |
| Restlessness | 2 (16.67) | 3 (23.08) | 5 (20) | 1.00 |
| Weight gain | 1 (8.33) | 4 (30.77) | 5 (20) | 0.32 |
| Constipation | 2 (16.67) | 2 (15.38) | 4 (16) | 1.00 |
| ***Decreased appetite*** | ***4 (33.33)*** | ***0 (0)*** | ***4 (16)*** | ***0.04*** |
| Drowsiness | 2 (16.67) | 1 (7.69) | 3 (12) | 0.59 |
| Diarrhea | 2 (16.67) | 1 (7.69) | 3 (12) | 0.59 |
| Headache | 1 (8.33) | 1 (7.69) | 2 (8) | 1.00 |
| Confused state | 1 (8.33) | 0 (0) | 1 (4) | 1.00 |
| Depression | 1 (8.33) | 0 (0) | 1 (4) | 1.00 |
| Decreased motor activity | 0 (0) | 1 (7.69) | 1 (4) | 1.00 |
| Blurred vision | 0 (0) | 1 (7.69) | 1 (4) | 1.00 |
| Dizziness | 1 (8.33) | 0 (0) | 1 (4) | 1.00 |
| Rash | 0 (0) | 1 (7.69) | 1 (4) | 1.00 |
| Weight loss | 1 (8.33) | 0 (0) | 1 (4) | 1.00 |
| Gastroesophageal reflux | 1 (8.33) | 0 (0) | 1 (4) | 1.00 |

**Supplemental Table 3: Incidence of Symptoms Elicited by the modified Dosage Record Treatment Emergent Symptom (MDOTES) scale during the supplement treatment.**

| **Treatment** | | | | | | | | | | | | |
| --- | --- | --- | --- | --- | --- | --- | --- | --- | --- | --- | --- | --- |
| **Symptom** | **Time** | | | | | | | | | | | |
|  | ***Week 0*** | | | ***Week 4*** | | | ***Week 8*** | | | ***Week 12*** | | |
|  | **1** | **2** | **3** | **1** | **2** | **3** | **1** | **2** | **3** | **1** | **2** | **3** |
| Confused State, (%) | 0 | 0 | 0 | 0 | 0 | 0 | 0 | 0 | 0 | 6.3 | 0 | 0 |
| Excitement/ agitation, (%) | 0 | 13 | 6.3 | 13 | 19 | 0 | 0 | 0 | 6.3 | 25 | 6.3 | 6.3 |
| Depression, (%) | 0 | 0 | 0 | 0 | 0 | 6.3 | 0 | 0 | 0 | 0 | 0 | 0 |
| Increased motor activity, (%) | 0 | 6.3 | 6.3 | 13 | 0 | 0 | 0 | 0 | 0 | 0 | 6.3 | 0 |
| Decreased motor activity, (%) | 0 | 6.3 | 0 | 0 | 0 | 0 | 0 | 0 | 0 | 0 | 0 | 0 |
| Insomnia, (%) | 13 | 0 | 0 | 0 | 6.3 | 0 | 0 | 0 | 0 | 0 | 0 | 0 |
| Drowsiness, (%) | 0 | 0 | 0 | 0 | 0 | 0 | 0 | 0 | 0 | 6.3 | 6.3 | 0 |
| Restlessness, (%) | 0 | 6.3 | 0 | 0 | 0 | 0 | 0 | 0 | 0 | 13 | 0 | 0 |
| Dry mouth/ excessive thirst, (%) | 6.3 | 0 | 0 | 6.3 | 0 | 0 | 6.3 | 0 | 0 | 6.3 | 0 | 0 |
| Nasal congestion, (%) | 0 | 0 | 0 | 6.3 | 0 | 0 | 0 | 0 | 0 | 25 | 0 | 0 |
| Blurred vision, (%) | 0 | 0 | 0 | 0 | 0 | 0 | 0 | 0 | 0 | 0 | 0 | 0 |
| Constipation, (%) | 6.3 | 0 | 0 | 0 | 6.3 | 6.3 | 0 | 6.3 | 0 | 0 | 0 | 0 |
| Diarrhea, (%) | 0 | 0 | 0 | 0 | 13 | 0 | 0 | 0 | 0 | 6.3 | 0 | 0 |
| Dizziness, (%) | 0 | 0 | 0 | 0 | 0 | 0 | 0 | 0 | 0 | 0 | 0 | 0 |
| Rash, (%) | 0 | 0 | 0 | 0 | 0 | 0 | 0 | 0 | 0 | 0 | 0 | 0 |
| Weight gain, (%) | 13 | 0 | 0 | 6.3 | 0 | 0 | 0 | 0 | 6.3 | 0 | 0 | 6.3 |
| Weight loss, (%) | 0 | 0 | 0 | 0 | 0 | 0 | 0 | 6.3 | 0 | 0 | 0 | 0 |
| Decreased appetite, (%) | 0 | 0 | 0 | 0 | 6.3 | 0 | 6.3 | 6.3 | 0 | 6.3 | 6.3 | 0 |
| Headache, (%) | 0 | 6.3 | 0 | 0 | 0 | 0 | 0 | 6.3 | 0 | 0 | 6.3 | 0 |
| Aggression, (%) | 13 | 6.3 | 0 | 0 | 6.3 | 0 | 0 | 6.3 | 6.3 | 6.3 | 6.3 | 0 |
| Gastroesophageal reflux, (%) | 0 | 0 | 0 | 0 | 6.3 | 0 | 0 | 0 | 0 | 0 | 0 | 0 |
| Emotional lability, (%) | 6.3 | 6.3 | 0 | 6.3 | 6.3 | 0 | 13 | 0 | 0 | 6.3 | 13 | 0 |
| Increased tantrums, (%) | 6.3 | 13 | 0 | 6.3 | 13 | 0 | 13 | 6.3 | 0 | 31 | 6.3 | 0 |

**Supplemental Table 4: Incidence of Symptoms Elicited by the modified Dosage Record Treatment Emergent Symptom (MDOTES) scale during the placebo treatment.**

| **Placebo** | | | | | | | | | | | | |
| --- | --- | --- | --- | --- | --- | --- | --- | --- | --- | --- | --- | --- |
| **Symptom** | **Time** | | | | | | | | | | | |
|  | ***Week 0*** | | | ***Week 4*** | | | **Week 8** | | | ***Week 12*** | | |
|  | **1** | **2** | **3** | **1** | **2** | **3** | **1** | **2** | **3** | **1** | **2** | **3** |
| Confused State, (%) | 6.3 | 0 | 0 | 0 | 0 | 0 | 0 | 0 | 0 | 0 | 0 | 0 |
| Excitement/ agitation, (%) | 25 | 0 | 0 | 13 | 0 | 0 | 0 | 13 | 0 | 6.3 | 25 | 6.3 |
| Depression, (%) | 0 | 0 | 0 | 0 | 0 | 0 | 0 | 0 | 0 | 0 | 0 | 0 |
| Increased motor activity, (%) | 0 | 6.3 | 0 | 0 | 0 | 0 | 0 | 6.3 | 0 | 6.3 | 13 | 6.3 |
| Decreased motor activity, (%) | 0 | 0 | 0 | 0 | 0 | 0 | 0 | 0 | 0 | 0 | 6.3 | 0 |
| Insomnia, (%) | 0 | 0 | 0 | 0 | 0 | 0 | 6.3 | 0 | 0 | 25 | 0 | 0 |
| Drowsiness, (%) | 6.3 | 6.3 | 0 | 0 | 0 | 0 | 0 | 0 | 0 | 6.3 | 0 | 0 |
| Restlessness, (%) | 13 | 0 | 0 | 6.3 | 0 | 0 | 0 | 0 | 0 | 6.3 | 6.3 | 0 |
| Dry mouth/ excessive thirst, (%) | 6.3 | 0 | 0 | 0 | 0 | 0 | 0 | 0 | 0 | 19 | 0 | 0 |
| Nasal congestion, (%) | 13 | 0 | 0 | 0 | 6.3 | 0 | 13 | 0 | 0 | 0 | 0 | 0 |
| Blurred vision, (%) | 0 | 0 | 0 | 0 | 0 | 0 | 0 | 0 | 0 | 6.3 | 0 | 0 |
| Constipation, (%) | 0 | 0 | 0 | 0 | 0 | 0 | 0 | 0 | 0 | 13 | 0 | 0 |
| Diarrhea, (%) | 6.3 | 0 | 0 | 0 | 6.3 | 0 | 0 | 0 | 0 | 0 | 0 | 0 |
| Dizziness, (%) | 0 | 0 | 0 | 0 | 0 | 0 | 0 | 0 | 0 | 6.3 | 0 | 0 |
| Rash, (%) | 0 | 0 | 0 | 0 | 0 | 0 | 6.3 | 0 | 0 | 0 | 0 | 0 |
| Weight gain, (%) | 0 | 0 | 0 | 6.3 | 0 | 0 | 6.3 | 0 | 0 | 19 | 0 | 0 |
| Weight loss, (%) | 0 | 0 | 0 | 0 | 0 | 0 | 0 | 0 | 0 | 0 | 0 | 0 |
| Decreased appetite, (%) | 6.3 | 6.3 | 0 | 0 | 0 | 0 | 0 | 0 | 0 | 0 | 0 | 0 |
| Headache, (%) | 0 | 0 | 0 | 0 | 0 | 0 | 0 | 0 | 0 | 0 | 6.3 | 0 |
| Aggression, (%) | 0 | 6.3 | 0 | 6.3 | 0 | 0 | 6.3 | 6.3 | 0 | 19 | 13 | 0 |
| Gastroesophageal reflux, (%) | 0 | 0 | 0 | 0 | 0 | 0 | 0 | 0 | 0 | 0 | 0 | 0 |
| Emotional lability, (%) | 6.3 | 6.3 | 0 | 6.3 | 6.3 | 0 | 0 | 6.3 | 0 | 6.3 | 13 | 0 |
| Increased tantrums, (%) | 13 | 6.3 | 0 | 6.3 | 6.3 | 0 | 13 | 0 | 0 | 6.3 | 19 | 0 |
